# Longitudinal wastewater-based surveillance of vancomycin-resistant Enterococci in tertiary-care hospitals

**DOI:** 10.1101/2025.01.16.25320689

**Authors:** Emily Au, Nicole Acosta, Barbara J. Waddell, Jangwoo Lee, Kristine Du, R. Benson Weyant, Maria Bautista, Janine McCalder, Jennifer Van Doorn, Kashtin Low, September Stefani, Gail Visser, Rhonda Clark, Johann Pitout, Joseph Kim, Bayan Missaghi, Oscar Larios, Jamil Kanji, Joseph Vayalumkal, Jenine Leal, Paul Westlund, Robert R. Quinn, Matthew James, Bonita Lee, Xiao-Li Pang, Bruce Dalton, Kevin Frankowski, Christine O’Grady, John M. Conly, Casey R.J. Hubert, Michael D. Parkins

## Abstract

**Summary:** Vancomycin-resistant *Enterococcus* (VRE) is an important cause of healthcare-associated infections. We adapted wastewater-based surveillance as a tool to longitudinally monitor VRE in hospitals through the detection of vancomycin resistance genes *vanA* and *vanB*.

**Methods:** Wastewater from four tertiary-care hospitals (three adult and one pediatric, totaling >2300 inpatient beds) and all three municipal wastewater treatment plants (WWTP) in Calgary, Canada (∼1.4 million) was sampled weekly (March to September 2022) and every other week (September 2022 to March 2023). Wastewater pellets were collected, DNA extracted, and *vanA* and *vanB* quantified by qPCR. *vanA* and *vanB* gene copies were assessed as raw (copies/mL) and normalized with three different fecal biomarkers – total bacterial 16S-rRNA, *Bacteroides* HF183 16S-rRNA, and human 18S-rRNA. Raw and normalized *vanA* and *vanB* abundance from each site was compared with clinically identified infections, vancomycin prescribing and hemodialysis services.

**Findings:** *vanA* abundance was up to1085-fold higher p<0.0001, Mann-Whitney) and *vanB* up to 32-fold higher (p<0.01, Mann-Whitney) in adult hospitals compared to an aggregate municipal signal and exhibited significantly greater variation. Strong correlations between each method of fecal normalization and raw-measured *vanA* and *vanB* were observed, and no analysis method proved superior (Spearman’s r=0.50–0.96, p<0.0001). *vanA* abundance was strongly correlated with hemodialysis provision (Spearman’s r=0.8357, p<0.0001) but not vancomycin prescribing.

**Interpretation:** Wastewater-based surveillance is a comprehensive, real-time tool capable of longitudinal hospital surveillance with the potential to transform the ability of infection control and antimicrobial stewardship programs to dynamically track, understand, and mitigate nosocomial antimicrobial-resistant pathogens.

## 1. Introduction

Vancomycin-resistant *Enterococcus* (VRE) is an opportunistic antimicrobial-resistant organism (ARO) that is frequently responsible for healthcare-associated infections(1). *Enterococcus faecium* is the most identified VRE in Canada (∼99%), with *Enterococcus faecalis* only rarely identified(1). Resistance to vancomycin in *E. faecium* is predominately conferred by the *vanA* gene, identified in 90.9% isolates, followed by *vanB*(1). VRE bacteremia is associated with an all-cause mortality rate of approximately 33% in Canada and places a significant burden the healthcare system(2, 3). Furthermore, VRE infection rates are on the rise. The Canadian Nosocomial Infection Surveillance Program (CNISP) reported a 72.2% increase in VRE bacteremia from 2016-2020(1, 4). From 2019-2022, healthcare-associated VRE bacteremia rates doubled from 0.27 to 0.47 per 10,000-patient days in Western Canada – disproportionately higher than the Canadian national average increase of 0.27-0.33(3). The Centers for Disease Control and Prevention (CDC) in the United States similarly reported a 14% increase in VRE hospital-associated infections from 2019-2020(5).

There are numerous factors that may contribute to increased risk of VRE colonization and infection in hospitals. Patient-level risk factors for VRE include but are not limited to length of stay, lack of isolation precautions, hemodialysis, surgery, transplantation, and antibiotic usage(6, 7). Hemodialysis patients are at a much greater risk of VRE colonization and infection due to their frequent hospitalizations and the selective pressures resulting from vancomycin’s frequent use owing to expedient dosing for suspected Gram-positive infections(8, 9). Hospital size is another risk factor for VRE infection, with the highest VRE bacteremia rates seen in larger adult hospitals(3, 7). These risk factors along with staffing shortages and lapses in non-respiratory infection prevention and control (IPC) measures and antimicrobial stewardship (ASP) services because of the COVID-19 pandemic may have inadvertently contributed to increased risk of VRE(5).

Despite the ongoing risk associated with VRE in hospitals, many institutions have discontinued VRE colonization surveillance owing to its substantial costs(10, 11), limiting our ability to understand contributing factors. Wastewater-based surveillance (WBS), however, may offer a unique approach for VRE and other ARO surveillance in hospitals. WBS is a platform technology that comprehensively and inclusively informs on the health of entire populations in a non-invasive and cost-effective manner, providing objective and actionable information(12). As an unbiased, objective, and actionable surveillance method, WBS has demonstrated itself as a powerful tool for public health. Leveraging a real-time hospital-based WBS network for SARS-CoV-2(13), we sought to understand the prevalence and determinants of VRE genes in wastewater of hospitals in Calgary, Alberta, Canada.

## 2. Methods and materials

### 2.1. Wastewater sampling

All inpatient care in the Calgary metropolitan area (population ∼1.4 million) and surrounding communities (total catchment 1.8 million) is provided by Alberta Health Services (AHS) though five tertiary care hospitals(Table 1 and Supplementary Figure 1). Hospital-1 and −2 (∼650 and ∼600 inpatient beds, respectively) are monitored by a single site. Hospital-3 is Calgary’s largest (∼1100 inpatient beds) and is regionally monitored by three non-overlapping collection sites exclusively capturing hospital wastewater. Hospital-3A monitors the smallest catchment including medical and surgical intensive care and surgical wards. Hospital-3B includes several hospital buildings and specifically includes the largest dedicated inpatient hemodialysis facilities. Hospital-3C includes the remainder of patient care units. Hospital-4 is a regional pediatric hospital (141 inpatient beds) that also utilizes zonal wastewater monitoring where inpatient and outpatient areas are captured by sites 4A and 4B (Supplementary Figure 1). Together these four facilities represent 90.2% of the region’s hospital beds.

**Table 1.**
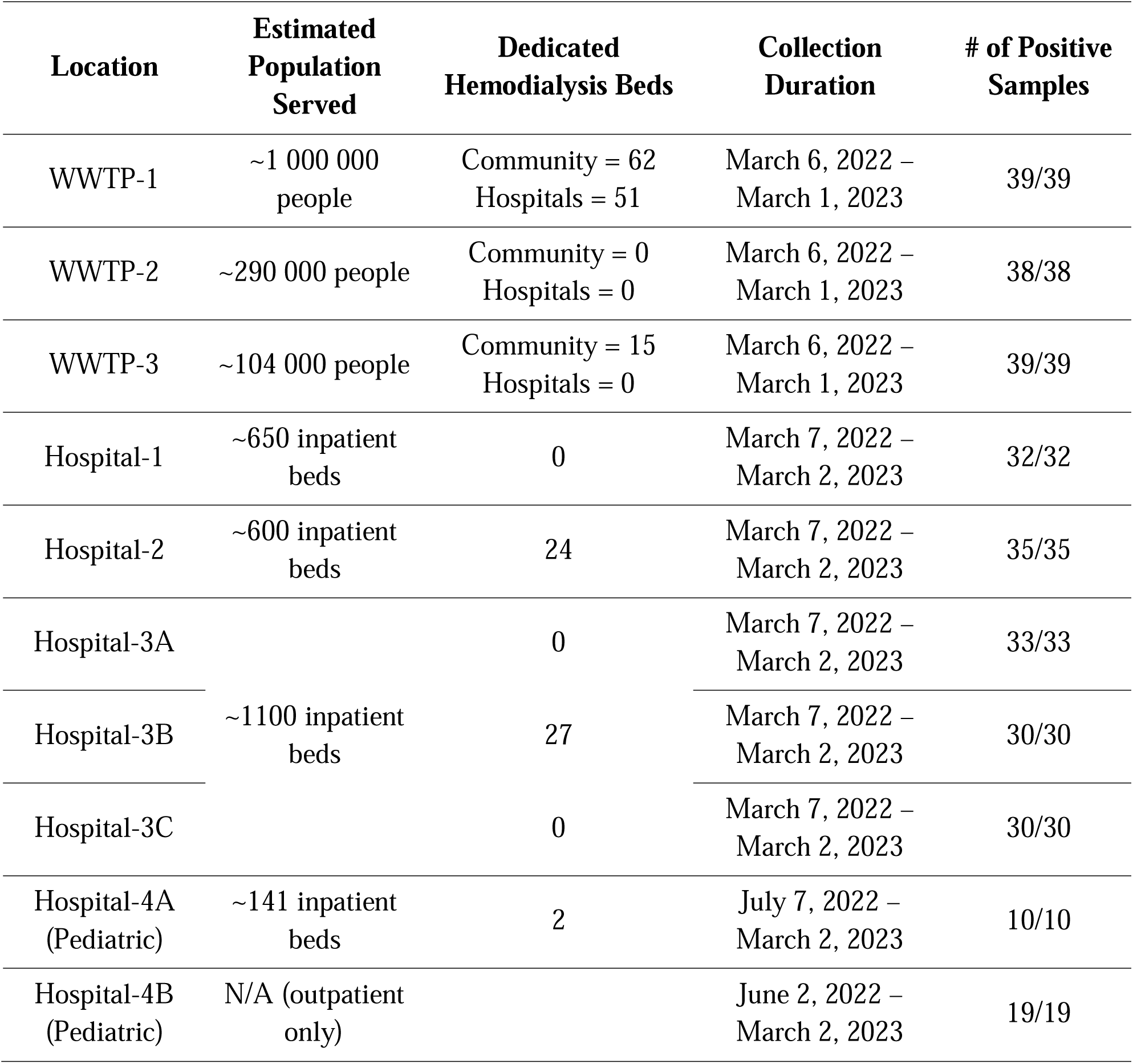
Calgary hospitals and wastewater treatment plants monitored for VRE *vanA* and *vanB* genes.

| Location | Estimated Population Served | Dedicated Hemodialysis Beds | Collection Duration | # of Positive Samples |
| --- | --- | --- | --- | --- |
| WWTP-1 | ~1 000 000 people | Community = 62<br>Hospitals = 51 | March 6, 2022 –<br>March 1, 2023 | 39/39 |
| WWTP-2 | ~290 000 people | Community = 0<br>Hospitals = 0 | March 6, 2022 –<br>March 1, 2023 | 38/38 |
| WWTP-3 | ~104 000 people | Community = 15<br>Hospitals = 0 | March 6, 2022 –<br>March 1, 2023 | 39/39 |
| Hospital-1 | ~650 inpatient beds | 0 | March 7, 2022 –<br>March 2, 2023 | 32/32 |
| Hospital-2 | ~600 inpatient beds | 24 | March 7, 2022 –<br>March 2, 2023 | 35/35 |
| Hospital-3A | ~1100 inpatient beds | 0 | March 7, 2022 –<br>March 2, 2023 | 33/33 |
| Hospital-3B |  | 27 | March 7, 2022 –<br>March 2, 2023 | 30/30 |
| Hospital-3C |  | 0 | March 7, 2022 –<br>March 2, 2023 | 30/30 |
| Hospital-4A<br>(Pediatric) | ~141 inpatient beds | 2 | July 7, 2022 –<br>March 2, 2023 | 10/10 |
| Hospital-4B<br>(Pediatric) | N/A (outpatient only) |  | June 2, 2022 –<br>March 2, 2023 | 19/19 |

Influent wastewater collected from the City of Calgary’s three wastewater treatment plants (WWTP) were used as a community reference for which to compare each hospital (Table 1)(14). Wastewater was collected weekly from March – September 7, 2022 and every second week from September 21, 2022 – March 2023. ISCO 6712 (Teledyne ISCO, USA) autosamplers were used to collect 10L of composite wastewater from each WWTP and Hospitals-1, −2, and −3A, −3B and −3C prior to entering municipal sewers (sampled at 15-minute intervals and pooled over 24 hours). CEC Analytics V1 (C.E.C. Analytics, Canada) autosampler was used to collect 2L of composite wastewater over 24 hours from within Hospital-4A and −4B plumbing. Equipoise between the two devices has been previously established (13). Samples were transported and stored at 4°C before immediate processing as previously described with minor modifications(15).

### 2.3. Quantitative PCR analysis

A multiplex qPCR assay was used to identify VRE gene determinants *vanA* and *vanB* in wastewater after extensive validation (see Supplementary Methods and Supplemental Tables 1-3). In addition to measuring raw *vanA* and *vanB* (copies/mL), variation in fecal bioburden between wastewater samples was accounted for with three normalization targets assessed by qPCR using a singleplex assay for total bacterial 16S rRNA and a multiplex *Bacteroides* HF183 16S and human 18S rRNA assay as alternative fecal biomarkers. In accord with the medical literature, our primary analysis utilized 16S rRNA normalization (16). To understand temporal other confounders, *vanA*/*vanB* gene copies were compared with wastewater-measured SARS-CoV-2 and influenza A RT-qPCR (see supplement).

### 2.4. Clinical data comparison

VRE colonization surveillance is no longer routinely performed in any AHS facilities(17). Accordingly, several indirect clinical sources were used as metadata comparators.

*i)* To assess VRE as a proportion of *E. faecium*-positive clinical isolates (*i.e.*, isolated from urine, blood, wound samples), we compared hospital and community samples collected across all of Calgary (AHS) between January-December 2022(18). *E. faecalis* was excluded as it accounted for <1% of all VRE.
*ii)* To assess the disproportionate burden of *E. faecium* and VRE in hospitals we compared their site-specific occurrence in hospitalized individuals and an aggregate community total (including all sites other than hospitals and emergency rooms) relative to two comparator pathogens responsible for similar infections stemming from the gastrointestinal and genitourinary systems: *Escherichia coli* and *Klebsiella pneumoniae* complex (18).
*iii)* As dialysis is among the most important VRE bacteremia risk factors(6, 8, 9), population- and facility-level hemodialysis (e.g., number of hemodialysis sessions and recipients/facility) was evaluated on a monthly basis as a function of *vanA*/*vanB* in hospital and community wastewater.
*iv)* Vancomycin prescribing as defined daily dosage was collected from the AHS Tableau Dashboard (not publicly available).

This project was approved by the Conjoint Regional Health Ethics Board (REB20-1252).

### 2.5. Statistical analyses

Non-parametric Mann-Whitney tests were performed to establish if there were differences between two datasets (*e.g*., between sites and targets). To compare more than two sets of data, a non-parametric Kruskal-Wallis test was performed. Differences in proportions among categorical data were assessed using the Fisher’s exact test for pairwise comparisons. Spearman’s rank correlations were used to observe correlated abundances between *vanA* and *vanB* gene copies, and to compare fecal normalization targets. Additionally, comparisons between population- and facility-level rates of hemodialysis and *vanA* gene abundances were conducted with Spearman’s rank correlations. A time-lagged cross-correlation function (CCF) analysis was used to compare vancomycin prescribing and *vanA* abundance across the three adult hospitals. CCF analysis was performed using R software version 4.4.1. A p<0.05 was the standard for statistical significance used in all analyses with GraphPad Prism (version9.0, Dotmatics, USA).

## 3. Results

### 3.1. VRE gene determinants are markedly enriched in hospital wastewater

Thirty-nine weeks of samples were collected. Successful collection was more likely to occur at well-equipped municipal WWTP (99%; 116/117) compared to samples collected from hospital manholes (Hospital-1, −2, −3A, -B, -C, 82%; 160/195) and in-building (Hospital-4, 37%; 29/78, p<0.001)(Table 1).

Significantly greater aggregate abundances of both *vanA* and *vanB* were observed in every adult hospital site (Hospitals-1 to −3A/B/C) compared to the aggregate city (p<0.0001, Mann-Whitney)(Figure 1). The greatest burden of *vanA* was identified from hospitals with the largest hemodialysis programs (see below). For example, Hospital-3 (and in particular −3B) which has 24% of Calgary’s dedicated dialysis bed capacity had *vanA* and *vanB* signals that were 1085-fold and 12-fold, respectively greater than the community (Figure 1, Table 2). In contrast to adult hospitals, lower abundances of *vanA* and only minimally increased levels of *vanB* were observed at the pediatric inpatient monitored site (Hospital-4A)(p<0.0001, Mann-Whitney). In the outpatient pediatric site (Hospital-4B), significantly lower median aggregate abundances of both *vanA* and *vanB* were seen (p<0.01 and p<0.0001, Mann-Whitney, respectively).

**Figure 1.**
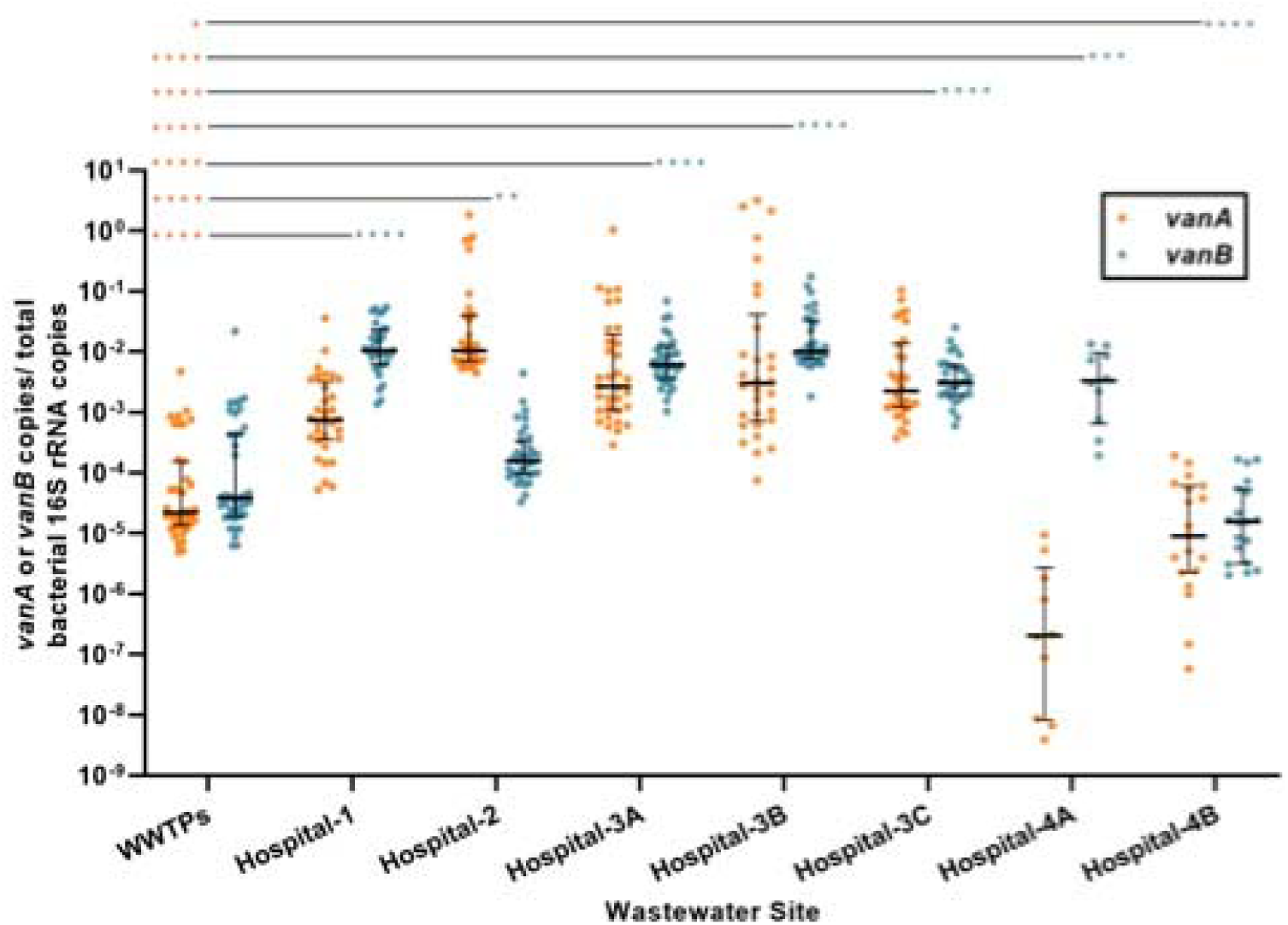
VRE gene determinants exists disproportionally in hospital wastewater. VRE *vanA* and *vanB* aggregate median abundances reported as a ratio of total bacterial 16S rRNA copies measured with qPCR at seven hospital wastewater sites (Adult sites: Hospital-1, −2, - 3A/B/C, and Pediatric sites: −4A/B) and a city wide metric incorporating all three Calgary wastewater treatment plants (WWTPs) from March 6, 2022 to March 2, 2023. Error bars represent interquartile ranges. Orange and blue asterisks respectively represent *vanA* and *vanB* p-values from Mann-Whitney comparisons between the combined WWTP signal and each hospital site (****p<0.0001, ***p<0.001, *p<0.01, *p<0.05).

**Table 2.** Hospital wastewater is markedly enriched for VRE. Mean fold increase in *vanA* and *vanB* (normalized for total bacterial 16S rRNA) in wastewater from each hospital site (Adult sites: Hospital-1, −2, −3A/B/C, and Pediatric sites: −4A/B) compared to a combined city wide metric incorporating all three WWTPs (Bonnybrook, Pine Creek, and Fish Creek) from March 6, 2022 to March 2, 2023.

| **Hospital** | ***vanA*** | ***vanB*** |
| --- | --- | --- |
| Hospital-1 | 9.48 | 19.26 |
| Hospital-2 | 507.18 | 0.44 |
| Hospital-3A | 173.03 | 2.35 |
| Hospital-3B | 1085.11 | 12.43 |
| Hospital-3C | 48.69 | 32.24 |
| Hospital-4A | 0.01 | 5.69 |
| Hospital-4B | 0.13 | 5.95 |

Mean total 16S rRNA-normalized *vanA* abundance exceeded *vanB* abundance across all sites except for Hospital-1 and Hospital-4 (pediatric)(Figure 1) – sites where VRE gene determinants were substantially lower than other hospitals. The greatest difference in proportion of *vanA* to *vanB* was seen in Hospital-2 (p<0.0001, Mann-Whitney). Within the community, there was no difference in the median aggregate abundance of 16S rRNA normalized *vanA* and *vanB* measured at the level of the WWTP. Despite large differences in gene copy numbers between *vanA* and *vanB,* Spearman’s rank correlations showed a moderate correlation between both VRE determinants at Hospital-1 (r=0.621, p=0.0002) and each Hospital-3 site (r=0.4956-0.7537, p<0.01, respectively)(Figure 2; only Hospital-3B shown).

**Figure 2.**
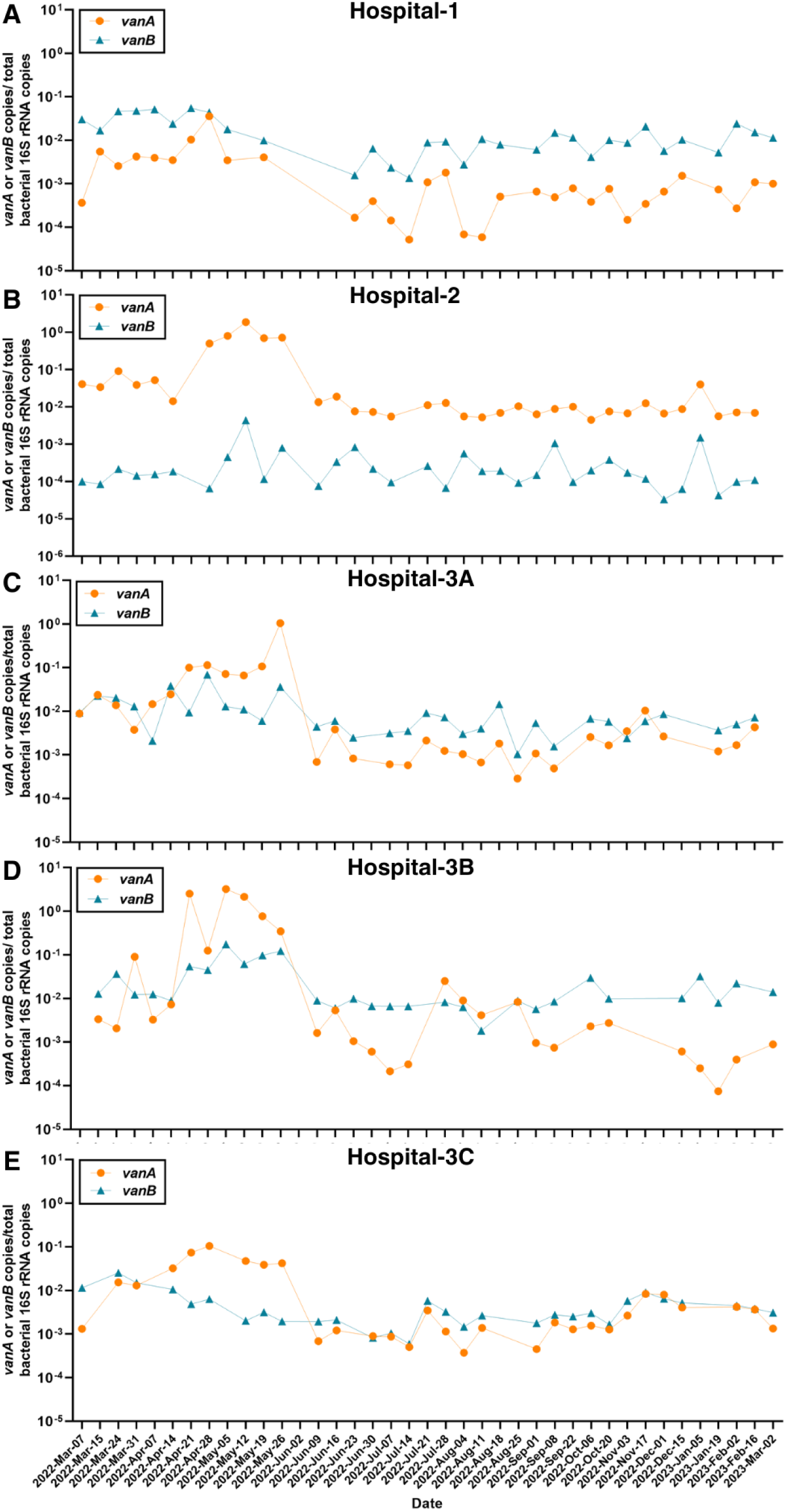
Longitudinal monitoring of VRE gene burden in wastewater from adult tertiary care hospitals demonstrates significant temporal variation. Mean *vanA* and *vanB* abundances normalized for total bacterial 16S rRNA measured from (A) Hospital-1, (B) Hospital-2, (C) Hospital-3A, (D) Hospital-3B, and (E) Hospital-3C wastewater as established by qPCR.

While a significant difference in 16S rRNA-normalized *vanA* and *vanB* abundances was observed between the three separate WWTPs used to assess city-wide burden of VRE gene determinants (*vanA*: p=0.0007 and *vanB*: p<0.0001, respectively), no difference in *vanA* was seen between WWTP-2 and −3 (Supplementary Figure 2). More importantly, the magnitude of difference between all three WWTPs is very small in comparison to the differences observed between the adult hospitals (Table 2).

### 3.2. Longitudinal fluctuations in *vanA* and *vanB* in wastewater

The coefficient of variation observed in hospital wastewater for *vanA* (total 16S rRNA-normalized) was greater at Hospital-1, −2, and −3A/B relative to a combined community signal, consistent with a dynamic burden of disease in a rapidly changing patient population (Figure 2). Similar observations were present with respect to *vanB*. The coefficient of variation of *vanB* was also lower than *vanA* for most sites (Figure 1). Every adult hospital site showed a period of increased *vanA* and *vanB* abundance from April to June 2022 (Figure 2). This trend was also observed in all three WWTPs (Supplementary Figure 3). Given the COVID-19 pandemic, SARS-CoV-2 abundance and COVID-19 infection was investigated as a potential contributor to increased VRE abundance. This period of elevated VRE gene determinants in hospital and community wastewater did not correlate with SARS-CoV-2 N1 or influenza A signals (https://covid-tracker.chi-csm.ca; hospital-site specific data not published)(data not shown). However, wastewater-detected *vanA* abundances correlated with the number of Calgary COVID-19-related hospitalizations in Hospital-2 (r=0.667, p=0.001) and Hospital-3B (r=0.777, p<0.0001) from March-August 2022 (Figure 3).

**Figure 3.**
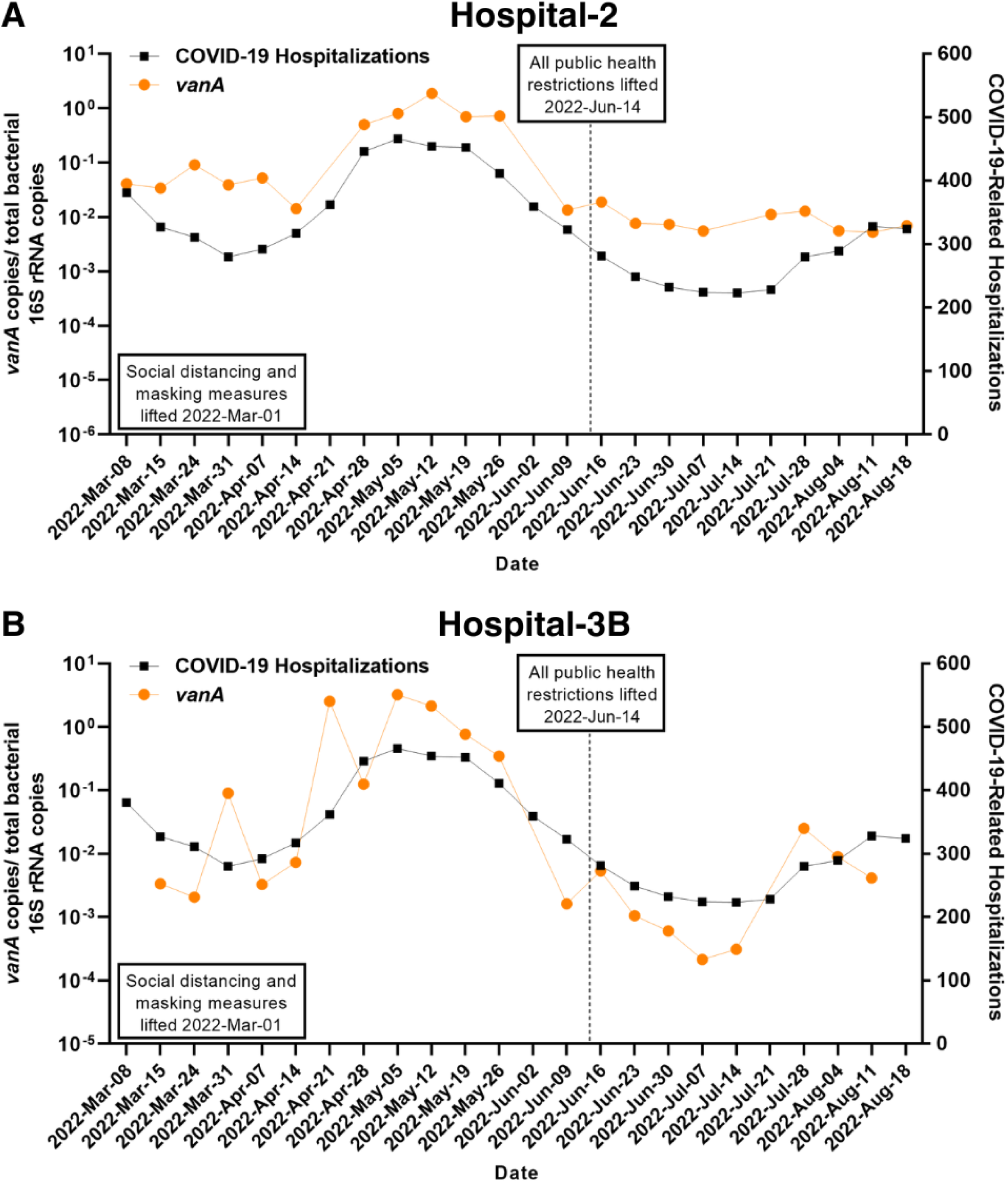
VRE infection and colonization in adult hospitals may associate with the burden of COVID-19 hospitalization. VRE *vanA* gene abundances in (A) Hospital-2 and (B) Hospital-3B measured with qPCR and assessed as a ratio of total bacterial 16S rRNA copies compared to total reported COVID-19-related hospitalizations from March 8, 2022 to August 18, 2022.

### 3.3. Effect of fecal normalization on VRE gene abundance measured in wastewater

Aggregate assessments of both *vanA* and *vanB* (Figures 1 and 4; Supplementary Figures 4-6) and longitudinal assessments of *vanA* (Supplementary Figure 7) from hospital wastewater demonstrates remarkably similar trends regardless of the three normalization methods. Figure 5, for example, compares Spearman’s rank correlations for each metric of *vanA* and *vanB* reporting for site Hospital-3B where all methods of normalization were significantly correlated with each other (*vanA*; r=0.90-96, p<0.0001 and *vanB*; r=0.50-69, p<0.0001). Similar results were observed for the other hospital sites (Supplementary Figure 8).

**Figure 4.**
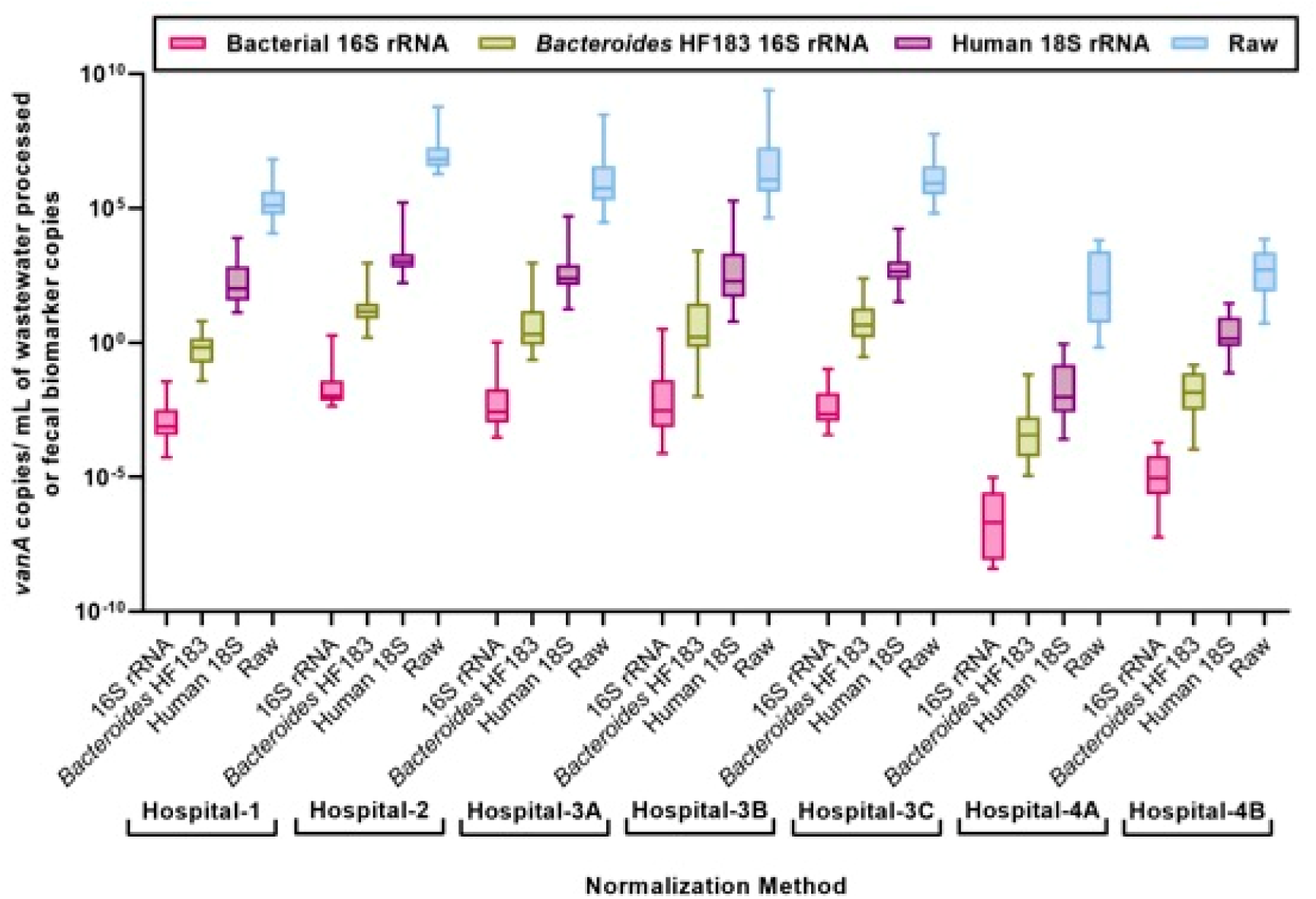
Normalization of *vanA* genes with different fecal biomarkers yields similar trends with aggregate assessment. Median aggregate *vanA* quantities assessed as raw values (copies per mL of wastewater) and ratios of different fecal biomarkers (total bacterial 16S rRNA, *Bacteroides* HF183 16S rRNA, and human 18S rRNA) as measured in Calgary tertiary care hospitals by qPCR from March 7, 2022 to March 2, 2023.

**Figure 5.**
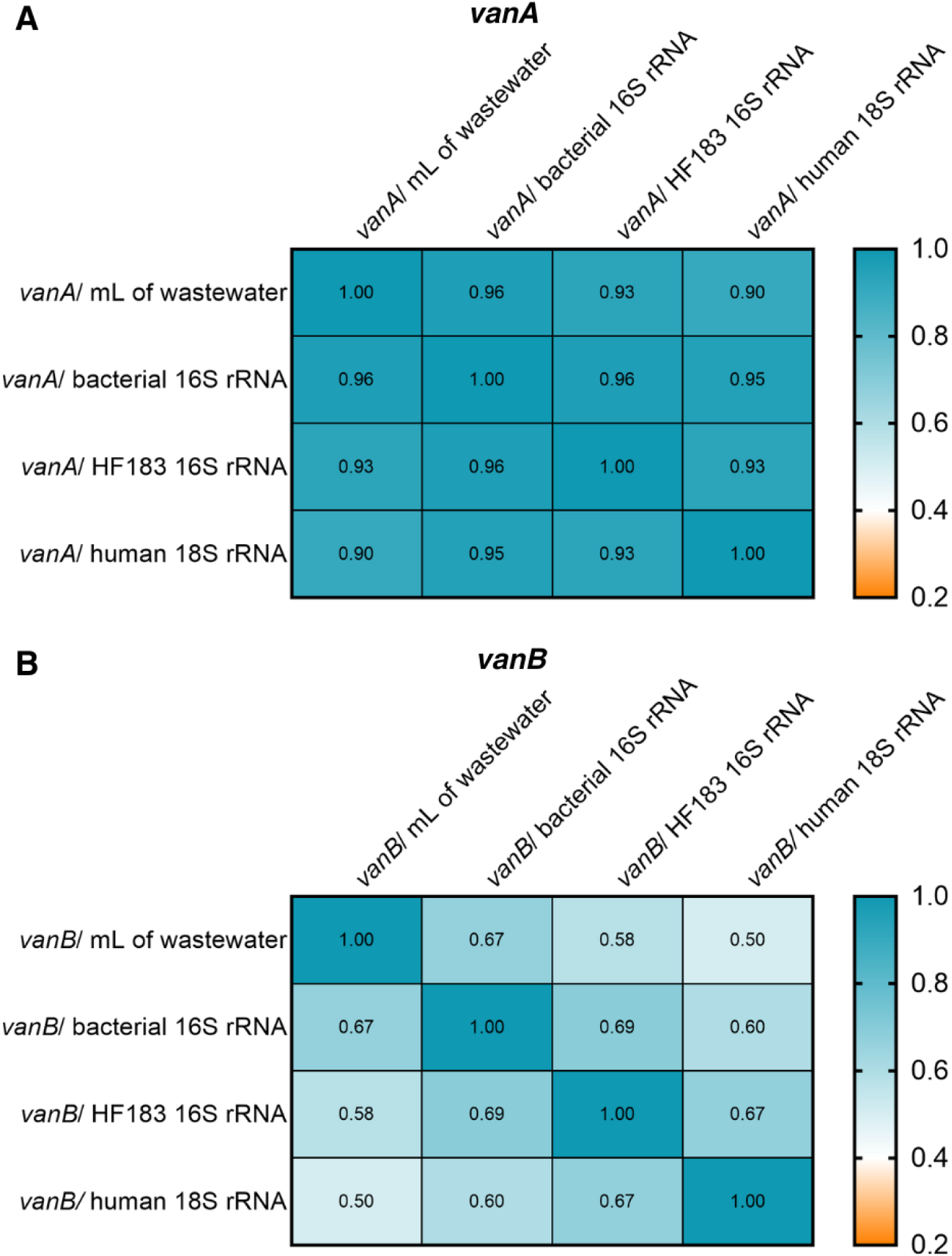
Quantities of *vanA* and *vanB* genes normalized with different fecal biomarker correlate strongly with each other and raw assessment. Spearman r correlations of (A) *vanA* and (B) *vanB* abundances in detected in Hospital-3B wastewater assessed as raw (copies per mL of wastewater) and normalized with three fecal biomarkers – total bacterial 16S rRNA, *Bacteroides* HF183 16S rRNA, and human 18S rRNA) demonstrating a high degree of correlation. (p<0.01).

### 3.5. VRE gene abundance in hospital wastewater correlates with clinical metadata

We confirmed that VRE infections occur disproportionally amongst those admitted to adult hospitals (*i.e.*, Hospitals-1, −2, −3) relative to that collected in the community (Supplementary Table 4). The observed relative risks of VRE as a proportion of *E. faecium* at each site relative to the community were Hospital-1: 2.72 (95% CI 1.40-5.27), p=0.003; Hospital-2: 2.51 (95% CI 1.40-4.53), p=0.009; Hospital-3: 2.51 (1.34-4.98), p<0.001; Hospital-4 (pediatric) could not be calculated because of too few VRE clinical isolates (Supplementary Table 5). Furthermore, relative to other pathogens responsible for similar infectious disease syndromes (*i.e.*, infections stemming from gastrointestinal or genitourinary systems) such as *Escherichia coli* and *Klebsiella pneumoniae* complex – VRE rates were markedly elevated in hospitalized populations (*i.e.,* the ratio of VRE to *E. coli* was increased 64-to 80-fold in hospitals relative to the community at large, whereas the ratio of VRE to *K. pneumoniae* complex was increased 25-to 34-fold).

Hemodialysis utilization in hospitals (inpatients and outpatients) and community clinics was compared to the abundance of *vanA* across all sites (Figure 6A). We observed a very strong correlation between total dialysis sessions performed each quarter and *vanA* abundance across each site (r=0.8357, p<0.0001, Spearman’s rank)(Figure 6B).

**Figure 6.**
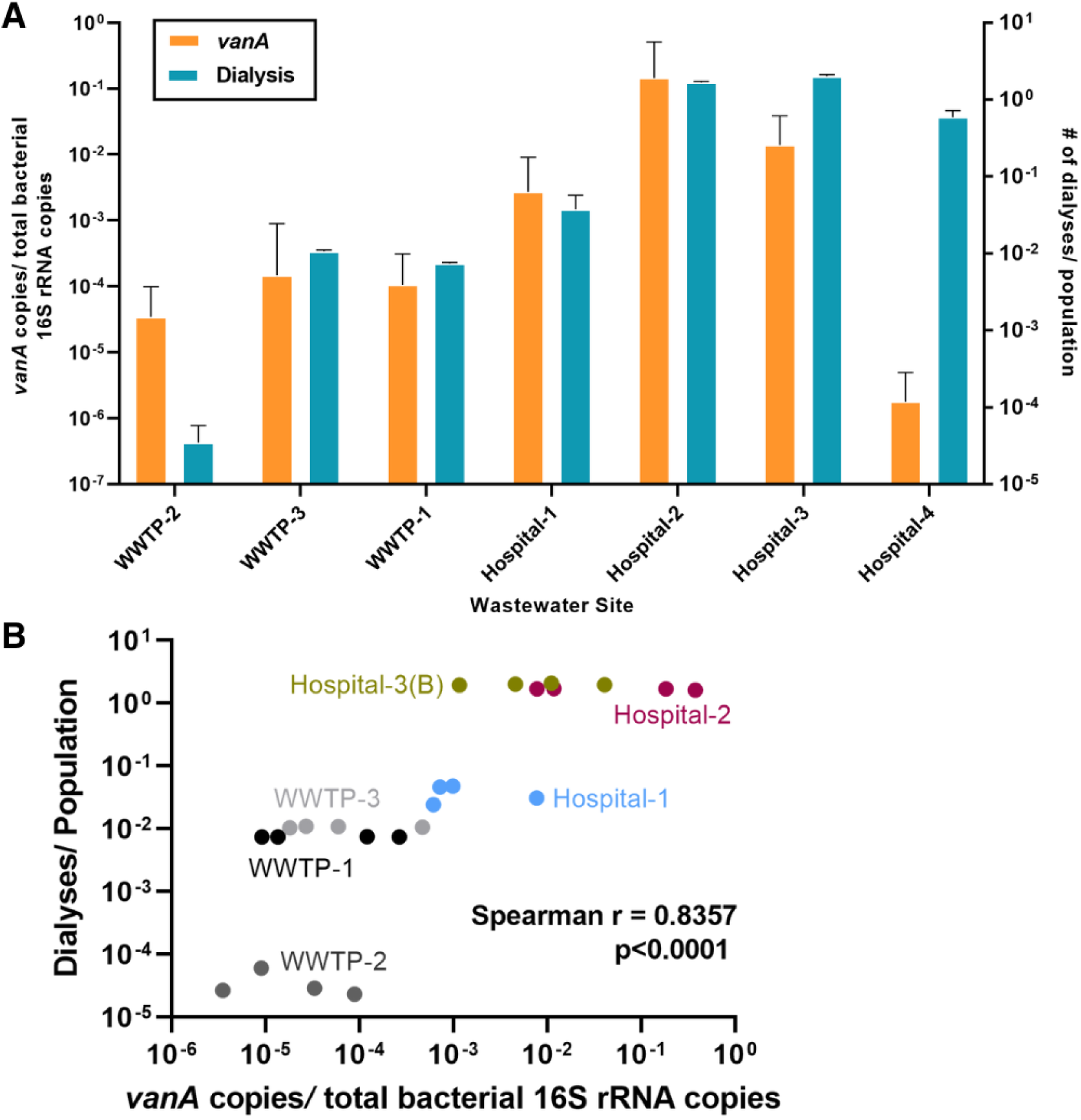
Dialysis is strongly correlated with VRE gene abundance. (A) Mean number of dialyses performed per population in community clinics captured by WWTP-1, −2, and −3, and Hospital-1, −2, −3, and −4 compared to the mean 16S rRNA-normalized *vanA* abundances measured with qPCR from the corresponding wastewater sites from April 2022 to March 2023. Only *vanA* abundances from wastewater sites Hospital-3B and Hospital-4A (inpatient) were used to represent Hospital-3 and Hospital-4, respectively. Error bars represent standard deviation. (B) The number of dialyses performed population strongly correlated with 16S rRNA-normalized VRE *vanA* abundances as measured with qPCR in hospital (Hospital-1, −2, and −3B) and community (WWTP-1, −2, and −3) wastewater. The proportion of dialyses and *vanA* gene copies were assessed as quarterly periods from April 2022 to March 2023 (i.e., April to June 2022, July to September 2022, October to December 2022, January to March 2023).

The highest rates of vancomycin prescribing occurred in those hospitals with higher wastewater measured *vanA,* where Hospital-3 had the greatest *vanA* abundance and vancomycin usage and Hospital-1 had the least vancomycin usage and detected *vanA* (Table 2, Supplementary Table 6). Aggregate *vanA* quantities and vancomycin defined daily doses for each adult hospital during this study were not significantly correlated (r=1.000, p=0.333, Spearman’s rank). Cross-correlation function analysis did not uncover strong leading relationships between vancomycin (DDD/100 PD) and *vanA* gene abundance across hospitals. A potential leading effect of oral vancomycin over *vanA* abundance was observed only in Hospital-2 (Supplementary Figure 9).

## 4. Discussion

Despite their importance, IPC and ASP programs – which are used by hospitals to prevent and mitigate the emergence of ARO– lack the tools necessary to perform nimble assessments of changing burden across hospitals. Furthermore, conventional ARO surveillance programs generally track AMR from those with clinically identified and confirmed infections.

Unfortunately, these infections are only a tiny fraction of the total population of individuals that are colonized by ARO, and any colonized individual is at risk both to develop (19) and transmit infections (20).

Hospital-based WBS may represent an important new tool to understand, control, and mitigate ARO in hospitals. Herein, we present for the first time a longitudinal hospital-based assessment of VRE – an important nosocomial pathogen. VRE was selected as the proof of concept pathogen on the basis of: *i)* it is responsible for a considerable burden of morbidity and mortality in hospital populations, *ii)* conventional surveillance programs have been trialed (and largely have since been abandoned) from which there is a robust literature to compare against, and *iii)* it can be accomplished with a limited number of molecular targets (*i.e.,* two AMR genes are predominately responsible) allowing for real-time monitoring capability. While protocols to screen admitted patients for VRE colonization have been developed(21), they have increasingly been abandoned primarily based on cost-efficiency concerns(22). Furthermore, even the most robust VRE screening programs generally assess for colonization only at hospital admission(21) despite acquisition risk increasing proportionate to the length of stay, thus inadequately quantifying and tracking the true prevalence of VRE(7). Novel technologies enabling cost-effective, serial surveillance is required to understand the dynamic burden of VRE. Hospital-based WBS programs, such as that described herein, may accomplish this mandate.

Using a combined WWTP signal as a community reference condition, we were able to confirm all adult tertiary-care hospital sites had markedly increased quantities of *vanA* and *vanB* as measured with qPCR. Within the pediatric Hospital-4, only *vanB* in the inpatient site (Hospital-4A) displayed significantly increased quantities relative to the community. Given the traditionally much lower risk of VRE infection in pediatric populations compared to adults, the overall VRE abundance detected in Hospital-4 is consistent with national surveillance of VRE which reported no pediatric VRE bacteremia episodes in 2021(1). In addition, the outpatient pediatric clinic populations (4B) – had even lower signals consistent with even lower antibiotic pressure and differing toileting patterns of visiting patients/families and staff.

VRE is primarily a healthcare-associated and nosocomial infection(1, 2), as illustrated by the fact that ∼90% of VRE bacteremia in Canada from 2017-2021 was healthcare-acquired(1). This epidemiology is reflected in our wastewater-derived findings, where *vanA* was detected (up to) 1084-fold greater quantities in hospitals than the community. Rates were highest in the largest hospital, consistent with published data correlating hospital size as a predictor of VRE (23). Furthermore, *vanA* abundance consistently exceeded *vanB* consistent with published studies of clinical isolates of VRE (1).

Much greater variation in VRE gene abundance was observed within and between individual hospital sites relative to the community. For example, WWTP-1 had a relatively smaller coefficient of variation for bacterial 16S rRNA-normalized *vanA* copies (197.2%). Given that the population (∼1 million) contributing to WWTP-1 does not change significantly from week to week and that VRE is not generally expected to be community-associated, *vanA* and *vanB* copies were not expected to fluctuate greatly each week. With the average length of hospital stay in Canada being 7.1 days (24), we can expect the hospital’s dynamic population to vary on a weekly basis and in turn, VRE prevalence – and this was observed. For example, Hospital-3B had greater fluctuation longitudinally (coefficient of variation = 262.5%), indicating much greater variation in *vanA* quantities over time within a hospital site compared to a community site.

The influence of hospital structure, function, and patient population was also assessed to understand the epidemiology of VRE gene determinants in wastewater. Hemodialysis is known to be among the strongest drivers of VRE prevalence(9) and not every hospital included in this study provided routine hemodialysis. Hospital-1 is the only hospital we monitored that does not conduct routine inpatient nor any outpatient hemodialysis and had mean aggregate *vanA* levels 53- and 114-fold lower than Hospital-2 and −3. Conversely, Hospital-3 had the greatest dialysis delivery capacity and recorded the highest mean aggregate abundance of both *vanA* and *vanB* at site 3B, which captures the inpatient and outpatient dialysis facilities. Hospital-2 closely followed Hospital-3 in dialysis provision and had the greatest aggregate abundance of *vanA* and *vanB.* In contrast, hemodialysis is performed at the pediatric Hospital-4 at a far lesser volume, with wastewater from this unit captured by the inpatient Hospital-4A sampler. Taken together, this study has shown that dynamic changes in *vanA* and *vanB*, both temporally and spatially, can be effectively tracked with WBS in hospital settings – an essential aspect for an ARO surveillance program to be actionable.

In contrast, we were not able to establish a strong correlation between vancomycin prescribing and *vanA* and *vanB* in wastewater. This is perhaps not surprising given that ASP interventions limiting vancomycin prescribing have not conclusively had the effect of reducing VRE occurrence in clinical studies(25) and that once established in hospital populations VRE is difficult to eliminate(26). Furthermore, many antibiotics well beyond vancomycin predispose individuals to VRE colonization and it may in fact be treatment duration that is more important than individual antibacterial agents (27).

As there is no current fecal normalization biomarker ‘gold standard’ in wastewater, we explored three different biomarkers – total bacterial 16S rRNA, human-specific *Bacteroides* HF183 16S rRNA, and human 18S rRNA. Among published AMR wastewater studies, bacterial 16S rRNA is generally used to normalize gene abundances (16). Each of the proposed normalization markers assessed has benefits and drawbacks (12). Here we demonstrate strong concordance of *vanA* and *vanB* abundance for raw measured wastewater and for the three fecal biomarkers used for normalization. However, until more information is available it may be advisable to use at least one fecal biomarker to correct for variation between wastewater samples, especially in proximal sewersheds (e.g., hospitals) with fewer contributors to the fecal signal.

Despite its potential for easy adaptation to clinical laboratory workflows and the ability for real-time data collection, there are a number of limitations that accompany qPCR-based WBS. While qPCR may possess greater sensitivity relative to other methods of analysis (e.g., metagenomics), it is only capable of identifying genes that are specifically targeted and cannot assess the entire resistome. Accordingly, only what is sought is identified. A more comprehensive approach to understand the entirety of the resistome, as well as to better characterize those organisms carrying *vanA*/*vanB* (*i.e.*, *E. faecium* versus *E. faecalis)* and the strain distribution would be required. As metagenomic technology evolves to become simpler, and more cost effective it may be possible for agnostic surveillance to perform targeted approaches such as *vanA* and *vanB* described herein. While *vanA* and *vanB* are the most prevalent VRE mechanisms and they associate with high-level adaptive resistance, other glycopeptide resistance mechanisms exist (*i.e*., the *vanC* gene is associated with intrinsic low-level resistance in rare human pathogens *Enterococcus casseliflavus* and *Enterococcus gallinarum –* although these are not reported, nor managed as VRE).

Limitations also exist within the wastewater sampling infrastructure. Despite the inclusivity of WBS, it is necessary to recognize that not all patients can feasibly be captured, as some of the sickest hospitalized patients may rely on continence aids that are disposed of as solid biologic waste. This limitation is particularly important to consider in pediatric wards (where most patients utilize diapers) and ICUs where many patients have rectal tubes/continence aids. We are also conscious that in outpatient environments of hospitals, toileting may be more infrequent for those with short visits(28). In such sites, there is likely insufficient fecal contributions for meaningful surveillance and results should be cautiously interpreted. We are similarly unable to differentiate between active fecal contributions with VRE and a persistent environmental contaminating signal. However, while VRE shed into hospital drains and pipes may contribute to biofilm and biofouling of pipes and create a background abundance, it should be relatively consistent and should not affect trends and fluctuations(29, 30). Additionally, a 24-hour composite wastewater sample collected on a weekly basis may not be truly representative of an entire week if there is increased shedding of VRE on days preceding and following sampling days. However, the potential for this bias is greater in a point-prevalence study than in a longitudinal surveillance study.

## 5. Conclusion

We have demonstrated WBS can be adapted to identify and quantify, and longitudinally monitor VRE resistance genes in hospital and community wastewater. VRE *vanA* and *vanB* genes were confirmed to exist at much greater abundance and exhibit significant variation over time in hospital wastewater relative to community wastewater, and correlate with multiple clinical metadata variables. Given the increasing incidence of AMR and healthcare-associated VRE infections, improved surveillance methods are essential for public health – and WBS programs have the potential to transform how IPC and ASP function, and even serve as a new primary outcome tool evaluating new intervention designed to reduce VRE colonization and infection in hospitals. Furthermore, WBS surveillance could be used as a cost-effective real-time monitoring tool that could trigger active population screening if changes in thresholds are observed.

## ACKNOWLEDGMENTS

The investigators are grateful to the staff of the City of Calgary Water Services, and Alberta Health Service Facilities Management for their support in providing wastewater samples for assessment. The authors gratefully acknowledge the front-line staff of the Alberta kidney programs who assisted in the collection of hemodialysis data. This work was supported by grants from the Canadian Institutes of Health Research, Alberta Health and Public Health Agency of Canada to MDP.

## AUTHOR CONTRIBUTIONS

EA prepared the original draft of the paper. EA, BJW, JL, KD, performed the molecular and genomic investigations and adapted the methodology. MB, JM, JvD, KL, SS, GV performed wastewater processing and cataloging. KF, CO’G, CRJH, MDP, BL and XP lead the Pan Alberta Wastewater network. EA, JP, JK, BM, OL, JK, JV, JL, RRQ, MJ, BL, XP, BD, JMC, and MDP collected and analyzed clinical information. EA, NA performed visualization. MDP led the study conceptualization. EA, NA, JL performed the formal analyses. All authors contributed to critical feedback on draft versions of the manuscript and approve the final version. RC led the project administration. MDP acquired funding and supervised the project and serves as guarantor of the work.

## DATA AVAILABILITY

Unlinked clinical metadata are available by request.

## References

1. CNISP. Healthcare-associated infections and antimicrobial resistance in Canadian acute care hospitals, 2017–2021. Can Commun Dis Rep. 2023;49(5):235-52.

2. Billington EO, Phang SH, Gregson DB, Pitout JD, Ross T, Church DL, et al. Incidence, risk factors, and outcomes for Enterococcus spp. blood stream infections: a population-based study. Int J Infect Dis. 2014;26:76–82.

3. Healthcare-Associated Infection (HAI) & Antimicrobial Resistant Organism (ARO) data [Internet]. Government of Canada. 2024 [cited March 11, 2024].

4. CNISP. Healthcare-associated infections and antimicrobial resistance in Canadian acute care hospitals, 2016-2020. Can Commun Dis Rep. 2022;48(7-8):308-24.

5. CDC. COVID-19: U.S. Impact on Antimicrobial Resistance, Special Report 2022. Report. Hyattsville, MD: 10.15620/cdc:117915; 2022.

6. Fossi Djembi L, Hodille E, Chomat-Jaboulay S, Coudrais S, De Santis N, Gardes S, et al. Factors associated with Vancomycin-resistant Enterococcus acquisition during a large outbreak. Journal of Infection and Public Health. 2017;10(2):185–90.

7. Johnstone J, Chen C, Rosella L, Adomako K, Policarpio ME, Lam F, et al. Patient- and hospital-level predictors of vancomycin-resistant Enterococcus (VRE) bacteremia in Ontario, Canada. American Journal of Infection Control. 2018;46(11):1266–71.

8. Zacharioudakis IM, Zervou FN, Ziakas PD, Rice LB, Mylonakis E. Vancomycin-resistant enterococci colonization among dialysis patients: a meta-analysis of prevalence, risk factors, and significance. Am J Kidney Dis. 2015;65(1):88–97.

9. D’Agata EMC, Green WK, Schulman G, Li H, Tang Y-W, Schaffner W. Vancomycin-Resistant Enterococci among Chronic Hemodialysis Patients: A Prospective Study of Acquisition. Clinical Infectious Diseases. 2001;32(1):23–9.

10. Pfister TR, Chow B, Shen Y, Ellison J, Bush K. The associated impact of standardized admission screening on vancomycin-resistant Enterococci bloodstream infections. Infection Control & Hospital Epidemiology. 2023;44(8):1289–93.

11. Cho SY, Kim HM, Chung DR, Choi JR, Lee MA, Huh HJ, et al. The impact of vancomycin-resistant Enterococcus (VRE) screening policy change on the incidence of healthcare-associated VRE bacteremia. Infect Control Hosp Epidemiol. 2022;43(5):603–8.

12. Parkins MD, Lee BE, Acosta N, Bautista M, Hubert CRJ, Hrudey SE, et al. Wastewater-based surveillance as a tool for public health action: SARS-CoV-2 and beyond. Clin Microbiol Rev. 2024;37(1):e0010322.

13. Acosta N, Bautista MA, Waddell BJ, Du K, McCalder J, Pradhan P, et al. Surveillance for SARS-CoV-2 and its variants in wastewater of tertiary care hospitals correlates with increasing case burden and outbreaks. Journal of Medical Virology. 2023;95(2).

14. Acosta N, Bautista MA, Waddell BJ, McCalder J, Beaudet AB, Man L, et al. Longitudinal SARS-CoV-2 RNA wastewater monitoring across a range of scales correlates with total and regional COVID-19 burden in a well-defined urban population. Water Research. 2022;220:118611.

15. Acosta N, Lee J, Bautista MA, Bhatnagar S, Waddell BJ, Au E, et al. Metagenomic analysis after selective culture enrichment of wastewater demonstrates increased burden of antibiotic resistant genes in hospitals relative to the community. medRxiv. 2023:2023.03.07.23286790.

16. Waseem H, Jameel S, Ali J, Saleem Ur Rehman H, Tauseef I, Farooq U, et al. Contributions and Challenges of High Throughput qPCR for Determining Antimicrobial Resistance in the Environment: A Critical Review. Molecules. 2019;24(1):163.

17. AHS. ARO Adult Antibiotic-resistant organisms (ARO) Acute Care Admission Screening FAQ. Canada 2017.

18. APL. Antibiograms: Calgary Zone 2022. 2024.

19. Alevizakos M, Gaitanidis A, Nasioudis D, Tori K, Flokas ME, Mylonakis E. Colonization With Vancomycin-Resistant Enterococci and Risk for Bloodstream Infection Among Patients With Malignancy: A Systematic Review and Meta-Analysis. Open Forum Infect Dis. 2017;4(1):ofw246.

20. Jackson SS, Harris AD, Magder LS, Stafford KA, Johnson JK, Miller LG, et al. Bacterial burden is associated with increased transmission to health care workers from patients colonized with vancomycin-resistant Enterococcus. Am J Infect Control. 2019;47(1):13–7.

21. OAHPP. Evidence review and revised recommendations for the control of vancomycin-resistant enterococci in all Ontario health care facilities. Toronto, Ontario; 2019.

22. MacDougall C, Johnstone J, Prematunge C, Adomako K, Nadolny E, Truong E, et al. Economic evaluation of vancomycin-resistant enterococci (VRE) control practices: a systematic review. J Hosp Infect. 2020;105(1):53–63.

23. Johnstone J, Garber G, Muller M. Health care-associated infections in Canadian hospitals: still a major problem. Canadian Medical Association Journal. 2019;191(36):E977.

24. CIHI. Hospital Stays in Canada Canada: Canadian Institute for Health Information; 2023 [Available from: https://www.cihi.ca/en/hospital-stays-in-canada.

25. de Bruin MA, Riley LW. Does vancomycin prescribing intervention affect vancomycin-resistant enterococcus infection and colonization in hospitals? A systematic review. BMC Infect Dis. 2007;7:24.

26. Kim WJ, Weinstein RA, Hayden MK. The changing molecular epidemiology and establishment of endemicity of vancomycin resistance in enterococci at one hospital over a 6-year period. J Infect Dis. 1999;179(1):163–71.

27. Gouliouris T, Warne B, Cartwright EJP, Bedford L, Weerasuriya CK, Raven KE, et al. Duration of exposure to multiple antibiotics is associated with increased risk of VRE bacteraemia: a nested case-control study. J Antimicrob Chemother. 2018;73(6):1692–9.

28. Heaton KW, Radvan J, Cripps H, Mountford RA, Braddon FE, Hughes AO. Defecation frequency and timing, and stool form in the general population: a prospective study. Gut. 1992;33(6):818.

29. Goudarzi M, Mobarez AM, Najar-Peerayeh S, Mirzaee M. Prevalence of biofilm formation and vancomycin-resistant genes among Enterococcus faecium isolated from clinical and environmental specimens in Lorestan hospitals. Iran J Microbiol. 2018;10(2):74–81.

30. Sukhum KV, Newcomer EP, Cass C, Wallace MA, Johnson C, Fine J, et al. Antibiotic-resistant organisms establish reservoirs in new hospital built environments and are related to patient blood infection isolates. Commun Med (Lond). 2022;2:62.

