## Supplementary Material file for "Longitudinal wastewater-based surveillance of vancomycin-resistant Enterococci in tertiary-care hospitals"

**1. Online Supplement**

**1.1 Supplementary Methods**

**1.1.1 Wastewater collection**

All hospitals sites monitored in this study are comprehensively and exclusively monitored by a through a wastewater sampling port (i.e. no community wastewater flows into these sites). Three plants serve the entire city of Calgary (Table 1 and Supplementary Figure 1) and each hospital’s wastewater is received at a specific municipal WWTP (Supplementary Figure 1). Wastewater signals from all three WWTPs were combined using daily wastewater flow rates and used as a reference condition for comparison with hospital sites (see below for details).

**1.1.2 *vanA*/*vanB* and fecal normalization biomarker qPCR Methods and Analysis**

The *vanA* and *vanB* multiplex qPCR assay adapted from He *et al.* (2020)(1) was validated using sequenced clinical VRE *E. faecium* and *E. faecalis* isolates and wastewater spiked with a VRE-containing mock community (see below for additional information).

To normalize the *vanA* and *vanB* gene copies with alternate fecal biomarkers *Bacteroides* HF183 and human 18S rRNA, *vanA* and *vanB* copies per mL of wastewater were assessed as a ratio of each fecal biomarker copies per mL of wastewater. qPCR testing on all samples were performed in triplicate, including a non-template water control for each plate. All qPCR assays were conducted with a QuantStudio^TM^ 6 Pro Real-Time PCR System (Applied Biosystems, USA).

Absolute abundance of all targets (*i.e.*, VRE *vanA* and *vanB,* and fecal biomarkers) was determined using target-specific standard curves (see below for additional details; Supplementary Tables 2-3). Calculation of gene copies per mL of wastewater was determined with the following calculation (E1):

$\text{Gene copies/mL of wastewater = C}\text{rxn}\text{ × 1/V}\text{rxn }\text{× V}\text{eluted }\text{× df}\text{DNA }\text{× df}\text{pellet }\text{× 1/V}\text{composite}$	(E1)

where C_rxn_ = number of gene copies per reaction exported from the QuantStudio^TM^ 6 Pro PCR System, V_rxn_ = volume of DNA added to each qPCR reaction (μL), V_eluted_ = total volume of DNA eluted during pellet extraction (μL), df_DNA_ = dilution factor of DNA, df_pellet_ = dilution factor of pellet, and V_composite_ = total volume of composite wastewater processed (mL).

Total bacterial 16S rRNA gene copies per reaction using *P. aeruginosa* PA01 genomic DNA as a standard curve were similarly converted to 16S rRNA copies per mL of wastewater with a modified calculation (2) (E2):

$\text{16S rRNA copies/mL of wastewater = E1 × }\left( \frac{\text{1 g}}{\text{1000}^{\text{3}}\text{ ng}} \right)\text{×}\left( \frac{\text{1 mol bp DNA}}{\text{600 g DNA}} \right)\text{×}\left( \frac{\text{6.023×}\text{10}^{\text{23}}\text{ bp}}{\text{1 mol bp}} \right)\text{×}\left( \frac{\text{PA01 16S rRNA copies}}{\text{PA01 genome size [bp]}} \right)$	(E2)

where *P. aeruginosa* PA01 has four 16S rRNA copies and a genome size of 6,463,464 bp(2).

To combine the signals from all three WWTPs as a sole community comparator for each of the gene targets, the following calculation was adapted (E3)(3):

$Combined WWTP signal per day=\left( C_{1}\times Q_{1} \right)+\left( C_{2}\times Q_{2} \right)+\left( C_{3}\times Q_{3} \right)$	(E3)

Where C = gene copies per mL of wastewater, Q = daily volumetric flow of wastewater (only available for WWTPs), and 1, 2, and 3 refer to each WWTP.

For each qPCR assay, a 10 μL reaction contained 5 μL of TaqMan^TM^ Fast Advanced Master Mix (Applied Biosystems, USA), 0.5 μL of primers (a mix of forward and reverse, per gene target), 0.5 μL of probe (per gene target), and 3 μL of template DNA (2.5 μL for normalization assays with 0.5 μL of UltraPure water). Primers and probes (Thermo Fisher Scientific^TM^, USA) sequences (Supplementary Table 1) were used with NCBI BLAST to determine the amplicon sequences for the VRE and *Bacteroides* HF183 16S rRNA/human 18S rRNA multiplex assays. The amplicon sequences as double-stranded gBlock DNA Gene Fragments (Integrated DNA Technologies^TM^, USA) were subsequently used as the standard curves for qPCR gene copy quantification (Supplementary Table 2). The gBlock standards were serially diluted to produce a standard curve with a large range of gene copies (Supplementary Table 3).

Thermocycling conditions were based on manufacturer recommendations for the TaqMan^TM^ Fast Advanced Master Mix (Applied Biosystems, USA) with slight modifications to optimize each set of qPCR targets. For the *vanA/vanB* multiplex, the only thermocycling modification was an increased cycle count of 50. While the total bacterial 16S rRNA used the standard number of cycles (40), the primer annealing temperature was increased from the standard 60°C to 62°C. Lastly, the normalization multiplex assay used 45 cycles, an annealing temperature of 55°C, and 3 seconds for denaturation and 30 seconds for annealing (standard times are 2 seconds and 20 seconds, respectively).

**1.1.3. Wastewater Spike Validation**

The *vanA/vanB* multiplex assay was originally validated by He *et al.* with 253 VRE and 90 non-VRE strains with 100% sensitive and specificity(1), and we further validated the assay with five previously whole genome sequenced local VRE clinical isolates (*E. faecium* and *E. faecalis*).

To validate that the qPCR assay was capable of accurately detecting differing quantities of VRE gene determinants in wastewater, samples were spiked with VRE and a mock community comprised of the following clinical isolates: vancomycin-resistant *Enterococcus faecium*, *C. difficile* NAP1, *Klebsiella pneumoniae bla*_NDM-1_, *Pseudomonas aeruginosa bla*_VIM-2_, and *Escherichia coli mcr-1.* All isolates were resuspended in a 0.85% saline solution to a McFarland 4 standard. To create the mock community, 50 μL of each isolate culture was pooled together and serially diluted by a factor of 100. Subsequently, 50 μL of 10^-2^, 10^-4^, and 10^-6^ dilutions were added to 200 μL of wastewater pellet. A wastewater pellet was also spiked with similar dilutions of exclusively VRE. A raw (non-spiked) pellet was also included as a control. The CFU/mL of VRE and mock community were determined using a 10^-6^ dilution grown on tryptic soy agar at 37°C overnight. Genomic DNA extractions were performed with the Qiagen PowerSoil Pro Kit (Qiagen, Germany) as previously described followed by qPCR to detect *vanA* and *vanB* to demonstrate respective decreases in detected VRE gene determinants with decreasing VRE spikes in wastewater.

**1.1.4. Assessing *vanA/vanB* in hospital and community wastewater as a function of SARS-CoV-2 and influenza A**

The abundance of SARS-CoV-2 and influenza A in hospital and community wastewater were determined in a separate study using similar molecular methods and analyses where a combined community wastewater signal was also utilized (4)(data not publicly available). SARS-CoV-2 N1 and influenza A *fluA* targets were assessed as copies per mL of wastewater processed. Reported cases of COVID-19 and influenza A in the Calgary Health Zone (including hospital-specific data) were retrieved from the Alberta Health Respiratory Virus Dashboard (5). Spearman rank correlation was used to compare *vanA/vanB* abundance with SARS-CoV-2/COVID-19 hospitalizations and influenza A.

**1.1.5. Hemodialysis Across the Calgary Health Zone**

WWTP-2 captured only a small fraction of hemodialysis, including only the regions smallest adult tertiary care hospital (310 beds) which was the only hospital not actively surveilled in our network (and does not have dedicated hemodialysis service and relies on outreach services). WWTP-3 captures a single large community hemodialysis clinic. In our analysis of the relationship between hemodialysis and VRE gene determinants, hemodialysis at each site was assessed as a function of individual runs, and patients receiving hemodialysis. Community hemodialysis sites were ascribed to their corresponding WWTP sewershed. WWTP-1 captured wastewater from Hospital-1, -2, -3, and -4, and four community outpatient hemodialysis clinics (Supplemental Figure 1). In the pediatric Hospital-4, all hemodialysis conducted was captured by the inpatient wastewater autosampler (Hospital-4A).

**1.1.6. Clinical Metadata Capture**

The monthly average total vancomycin defined daily dose (DDD) from March 2022 to February 2023 was assessed to compare vancomycin prescribing volume between the three adult hospitals. Oral and parenteral vancomycin DDD per 100 patient days (PD) for defined hospital populations corresponding to the wastewater catchments were obtained from Alberta Health Services Pharmacy Services. Cross-correlation function analysis was performed to compare oral, parenteral and total vancomycin usage (DDD/100 PD) to *vanA* abundance (16S rRNA-normalized) in all adult hospitals (Hospital-1/2/3A-C). We were unable to access community vancomycin prescribing.

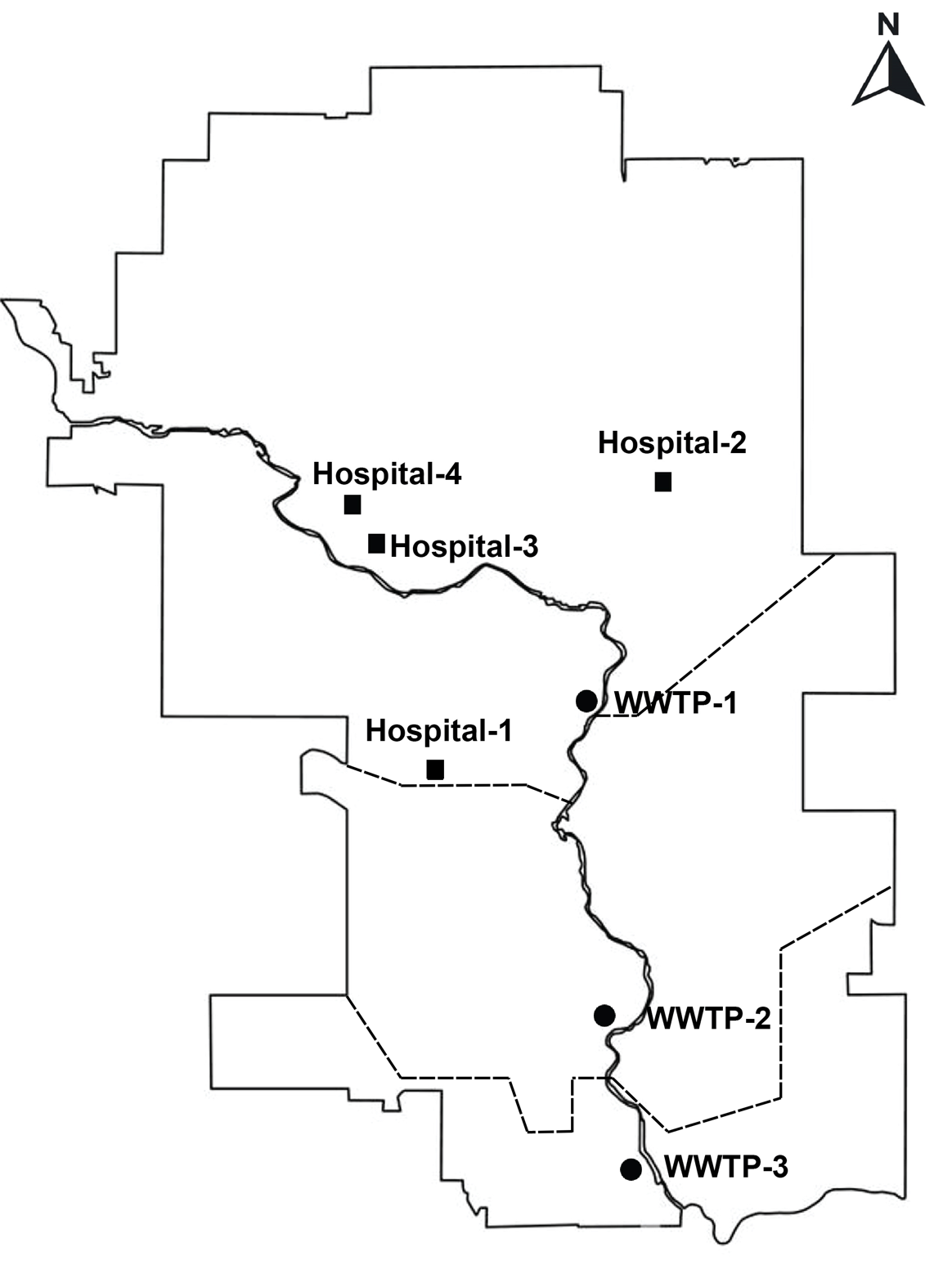

**Supplementary Figure 1.** Map of the City of Calgary detailing hospital and wastewater treatment plant (WWTP) locations. Dashed lines represent the boundaries of the areas captured by each WWTP.

**Supplementary Table 1.** Primers and probes for VRE and fecal biomarker detection with qPCR.

| **Vancomycin-resistant enterococci** | | | |
| --- | --- | --- | --- |
| **Target** | **Primers (5’ → 3’)** | **Probe (5’ → 3’)** | **Source** |
| *vanA* | F: GCCGGAAAAAGGCTCTGAA  R: TTTTTTGCCGTTTCCTGTATCC | FAM/CGCAGTTA  TAACCGTTCCCG  CAGACC/MGBNFQ | He *et al.* (2020) (1) |
| *vanB* | F: GATTTGATTGTCGGCGAAGTG  R: TCCTGATGGATGCGGAAGA | VIC/TCAAATCC  GGCTGAGCCACG  GT/MGBNFQ | He *et al.* (2020) (1) |
| **Fecal normalization biomarkers** | | | |
| **Target** | **Primers (5’ → 3’)** | **Probe (5’ → 3’)** | **Source** |
| Total bacterial 16S rRNA | F: TCCTACGGGAGGCAGCAGT  R: GGACTACCAGGGTATCTAATCCTGTT | FAM/CGTATTAC  CGCGGCTGCTG  GCAC/MGBNFQ | Nadkarni  *et al.* (2002) (6) |
| *Bacteroides* HF183 16S rRNA | F: ATCATGAGTTCACATGTCCG  R: CTTCCTCTCAGAACCCCTATCC | VIC/ATCGTTGA  CTAGGTGGGCCG  TTAC/MGBNFQ | Greenwald  *et al.* (2021) (7) |
| Human 18S rRNA | F: GGTTCCTTTGGTCGCTCGCT  R: GGGCTGACCGGGTTGGTTTT | FAM/AGAGCTAA  TACATGCCGAC  GGGC/MGBNFQ | Greenwald  *et al.* (2021) (7) |

FAM and VIC = probe dyes, MGBNFQ = probe quencher

**Supplementary Table 2.** gBlocks for VRE gene determinants and fecal biomarker detection with qPCR.

| **qPCR assay** | **gBlock amplicon sequence** | **Amplicon size** |
| --- | --- | --- |
| *vanA*/*vanB* | TATTCATCAGGAAGTCGAGCCGGAAAAAGGCTCTGAAAA  CGCAGTTATAACCGTTCCCGCAGACCTTTCAGCAGAGGA  GCGAGGACGGATACAGGAAACGGCAAAAAAAATATATAA  AGCGCTCGGCTGGGTCATGGGGAACGAGGATGATTTGAT  TGTCGGCGAAGTGGATCAAATCCGGCTGAGCCACGGTAT  CTTCCGCATCCATCAGGAAAACGAGCCGG | 224 bp |
| Total bacterial 16S rRNA | N/A – *P. aeruginosa* PA01 genomic DNA | N/A |
| *Bacteroides* HF183 16S rRNA/  human 18S rRNA | CATTAAATCAGTTATGGTTCCTTTGGTCGCTCGCTCCTC  TCCTACTTGGATAACTGTGGTAATTCTAGAGCTAATACA  TGCCGACGGGCGCTGACCCCCTTCGCGGGGGGGATGCGT  GCATTTATCAGATCAAAACCAACCCGGTCAGCCCGATTA  ATCCAGGATGGGATCATGAGTTCACATGTCCGCATGATT  AAAGGTATTTTCCGGTAGACGATGGGGATGCGTTCCATT  AGATAGTAGGCGGGGTAACGGCCCACCTAGTCAACGATG  GATAGGGGTTCTGAGAGGAAGGTCCCCCACATTG | 307 bp |

**Supplementary Table 3.** VRE and fecal normalization biomarker qPCR assay details.

| **Vancomycin-Resistant Enterococci** | | | | |
| --- | --- | --- | --- | --- |
| **Target** | **qPCR Efficiency (%)** | **Master Mix Content**  **(10 μL Total Reaction Volume)** | **qPCR Standard Curve Range** | **DNA Dilution Factor** |
| *vanA* | 90.6-95.3 | TaqMan^TM^ Fast Advanced master mix (5 μL)  *vanA* F+R primer mix (8000 nM, 0.5 μL)  *vanA* probe (4000 nM, 0.5 μL)  *vanB* F+R primer mix (8000 nM, 0.5 μL)  *vanB* probe (4000 nM, 0.5 μL)  Template DNA (3 μL) | 300 000 000-300 copies  (9 serial dilutions by a factor of 10) | 1:15 |
| *vanB* | 89.0-90.2 |  |  |  |
| **Fecal Normalization Biomarkers** | | | | |
| **Target** | **qPCR Efficiency (%)** | **Master Mix Content**  **(10 μL Total Reaction Volume)** | **qPCR Standard Curve Range** | **DNA Dilution Factor** |
| Total bacterial 16S rRNA | 92.8-99.6 | TaqMan^TM^ Fast Advanced master mix (5 μL)  16S F+R primer mix (8000 nM, 0.5 μL)  16S probe (2000 nM, 0.5 μL)  UltraPure H_2_O (1.5 μL)  Template DNA (2.5 μL) | 62.5-0.00016 copies  (9 serial dilutions by a factor of 5) | 1:15 |
| *Bacteroides* HF183 16S rRNA | 91.5-94.6 | TaqMan^TM^ Fast Advanced master mix (5 μL)  HF183 F+R primer mix (8000 nM, 0.5 μL)  HF183 probe (4000 nM, 0.5 μL)  18S F+R primer mix (8000 nM, 0.5 μL)  18S probe (4000 nM, 0.5 μL)  UltraPure H_2_O (0.5 μL)  Template DNA (2.5 μL) | 250 000 000-2.5 copies  (9 serial dilutions by a factor of 10) | 1:15 |
| Human 18S rRNA | 91.5-96.6 |  |  |  |

**1.2 Supplementary Results**

**1.2.1. Wastewater Spike Validation**

The *vanA/* *vanB* qPCR assay has previously been validated by He *et al.* (2020) with VRE clinical isolates responsible for causing disease (100% sensitivity and 100% specificity) (1), we further confirmed the assay validity with sequenced local VRE clinical isolates. In these isolates, the *vanA* and *vanB* genes identified with the qPCR assay were concordant with our sequencing results. Additionally, a panel of non-Enterococcal species including multiple human pathogens were used as negative controls. Additional studies using spiked wastewater with dilutions of VRE *E. faecium* (*vanA^+^*/*vanB*^-^) and a mock community containing VRE among other Gram-negative and Gram-positive clinical isolates demonstrated respective decreases in the quantity of *vanA* detected from the wastewater with qPCR (data not shown). As such, this *vanA*/*vanB* multiplex was adequately adapted to detect VRE in complex wastewater samples.

**1.2.2. Hemodialysis Utilization**

Among the hospitals, Hospital-3 has the greatest dialysis capacity (24% of the entire city) and conducted significantly more dialysis than Hospital-2 (p<0.0001, Mann-Whitney). Hospital-2 conducted significantly more dialysis than Hospital-1 (p<0.0001, Mann-Whitney), which does not possess a dedicated hemodialysis service and any performed there is on an as needed basis.

Only adult hospitals were included in statistical comparisons assessing *vanA* relative to the proportion of dialysis runs as the pediatric hospital had an insufficient number of wastewater samples.

**1.2.3. VRE Gene Determinant Abundances in Hospital and Community Wastewater**

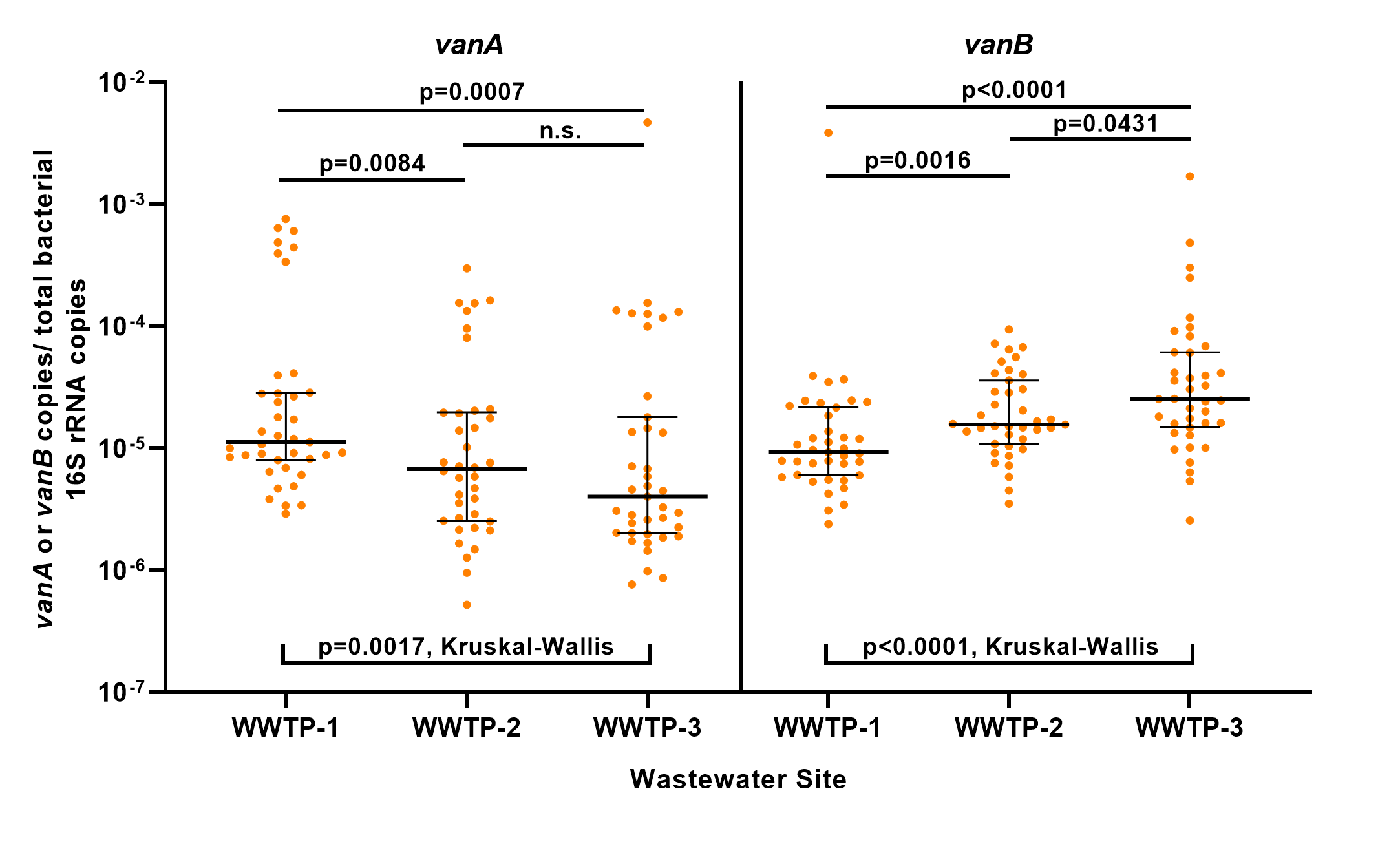

**Supplementary Figure 2.** Median aggregate abundance of *vanA* and *vanB* as a ratio of total bacterial 16S rRNA copies as measured by qPCR in three Calgary wastewater treatment plants (WWTP) from March 6, 2022 to March 1, 2023. Error bars represent interquartile ranges. Mann-Whitney (top) and Kruskal-Wallis (bottom) comparisons provided for site-to-site comparisons (n.s. = not significant).

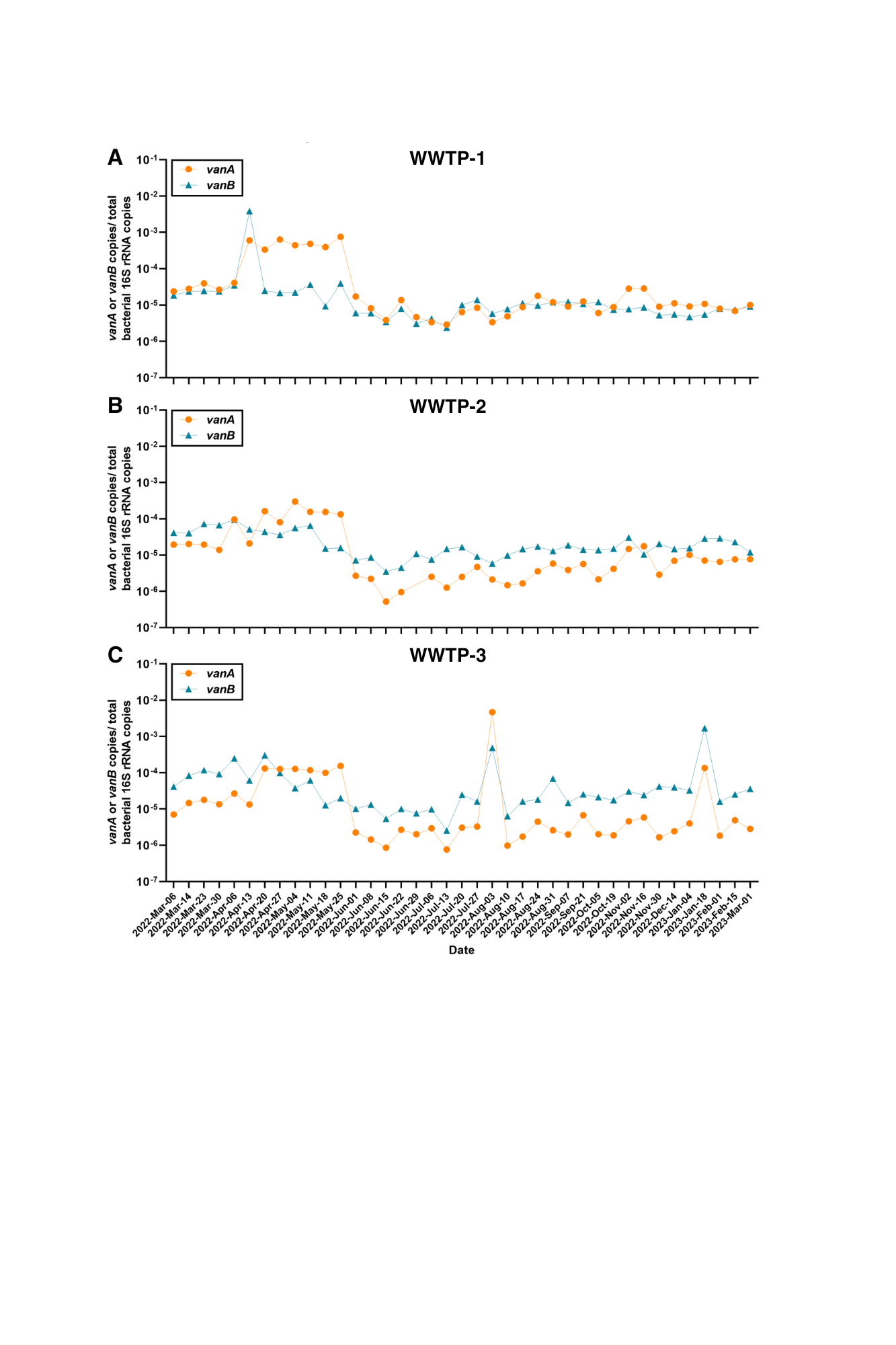

**Supplementary Figure 3. Longitudinal analysis of VRE gene burden in community wastewater from municipal wastewater treatment plants (WWTPs).** Mean *vanA* and *vanB* abundances normalized for total bacterial 16S rRNA measured from (A) Bonnybrook WWTP (WWTP-1), (B) Pine Creek WWTP (WWTP-2), and (C) Fish Creek WWTP (WWTP-3) from March 6, 2022 to March 1, 2023 as established by qPCR.

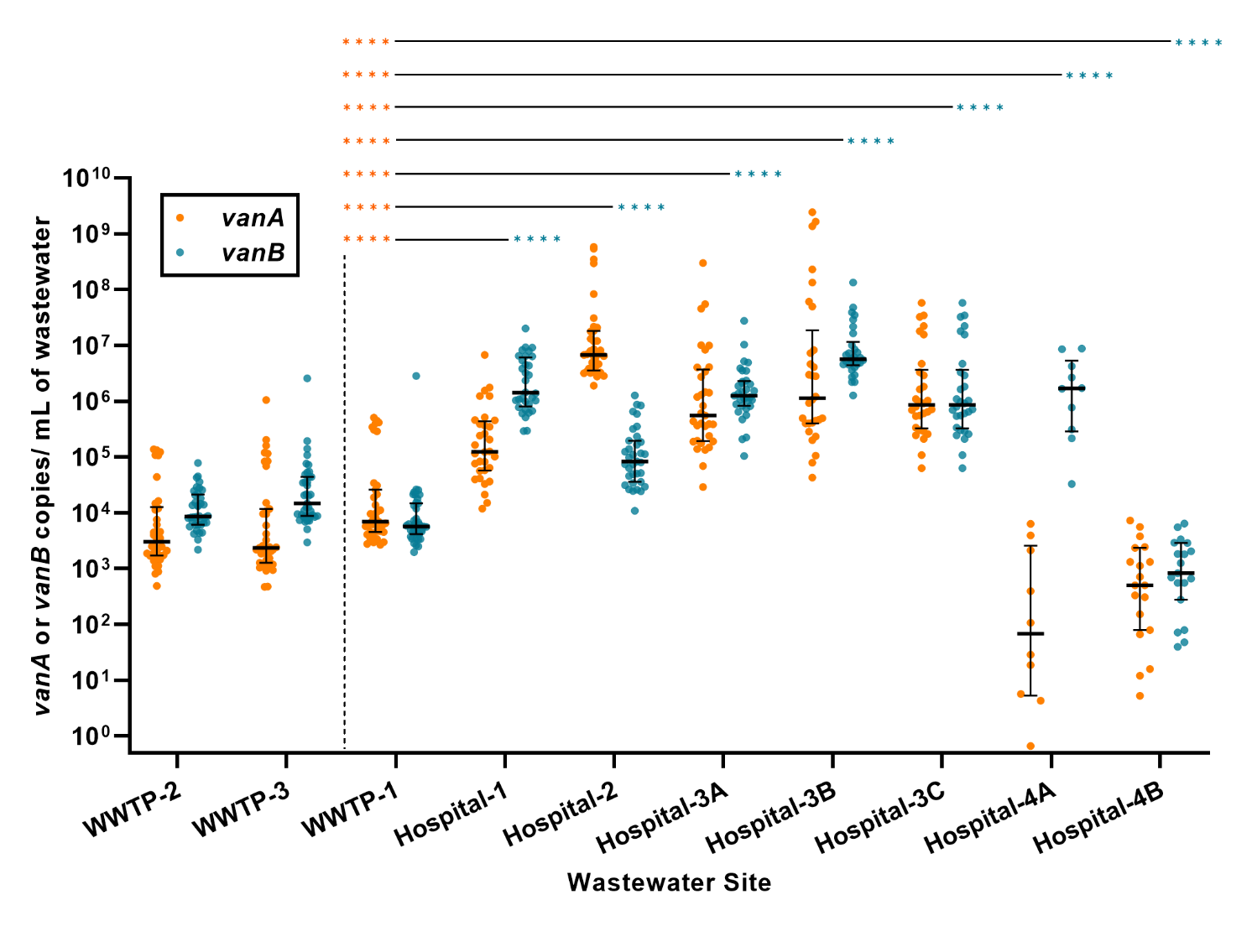

**Supplementary Figure 4.** VRE *vanA* and *vanB* aggregate median abundances assessed as raw copies per mL of wastewater processed, measured using qPCR from three WWTPs and seven hospital wastewater sites (Hospital-1, -2, -3A/B/C, -4A/B) sampled from March 6, 2022 to March 2, 2023. Error bars represent interquartile ranges. Orange and blue asterisks respectively represent *vanA* and *vanB* p-values from Mann-Whitney comparisons between WWTP-1 (the largest) and each hospital site (****p<0.0001). A combined municipal signal using raw copies per mL cannot be assessed on account it must be normalized by wastewater flow rates (m^3^/day). Additional WWTP data included to demonstrate similarity to WWTP-1.

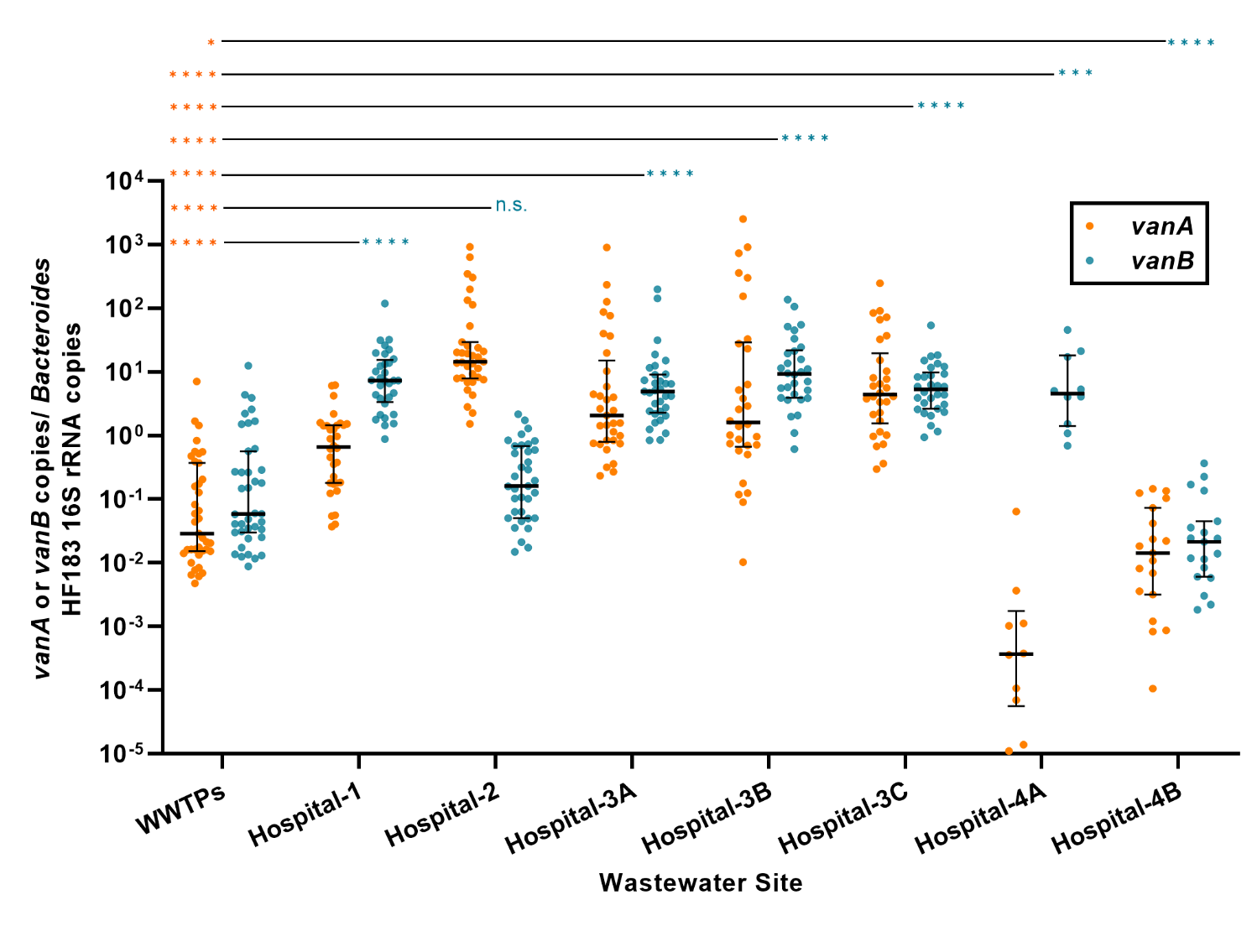

**Supplementary Figure 5.** VRE *vanA* and *vanB* aggregate median abundances as a ratio of *Bacteroides* HF183 16S rRNA copies measured using qPCR from three WWTPs combined and seven hospital wastewater sites (Hospital-1, -2, -3A/B/C, -4A/B) sampled from March 6, 2022 to March 2, 2023. Error bars represent interquartile ranges. Orange and blue asterisks respectively represent *vanA* and *vanB* p-values from Mann-Whitney comparisons between the combined WWTP site and each hospital site (****p<0.0001, ***p<0.001, *p<0.05, n.s.=not significant).

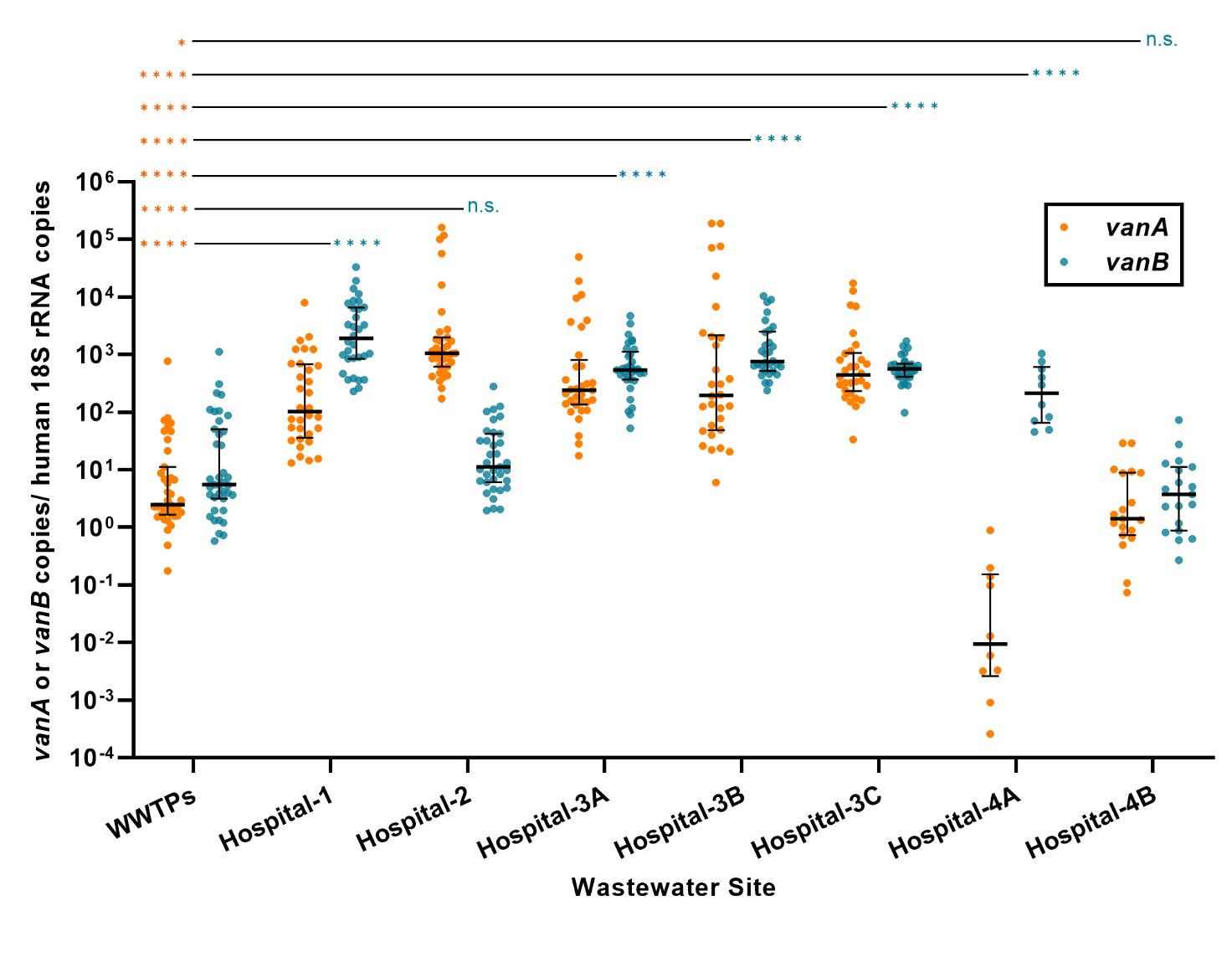

**Supplementary Figure 6.** VRE *vanA* and *vanB* aggregate median abundances as a ratio of human 18S rRNA copies measured using qPCR from three WWTPs combined and seven hospital wastewater sites (Hospital-1, -2, -3A/B/C, -4A/B) sampled from March 6, 2022 to March 2, 2023. Error bars represent interquartile ranges. Orange and blue asterisks respectively represent *vanA* and *vanB* p-values from Mann-Whitney comparisons between the combined WWTP site and each hospital site (****p<0.0001, **p<0.01, *p<0.05, n.s.=not significant).

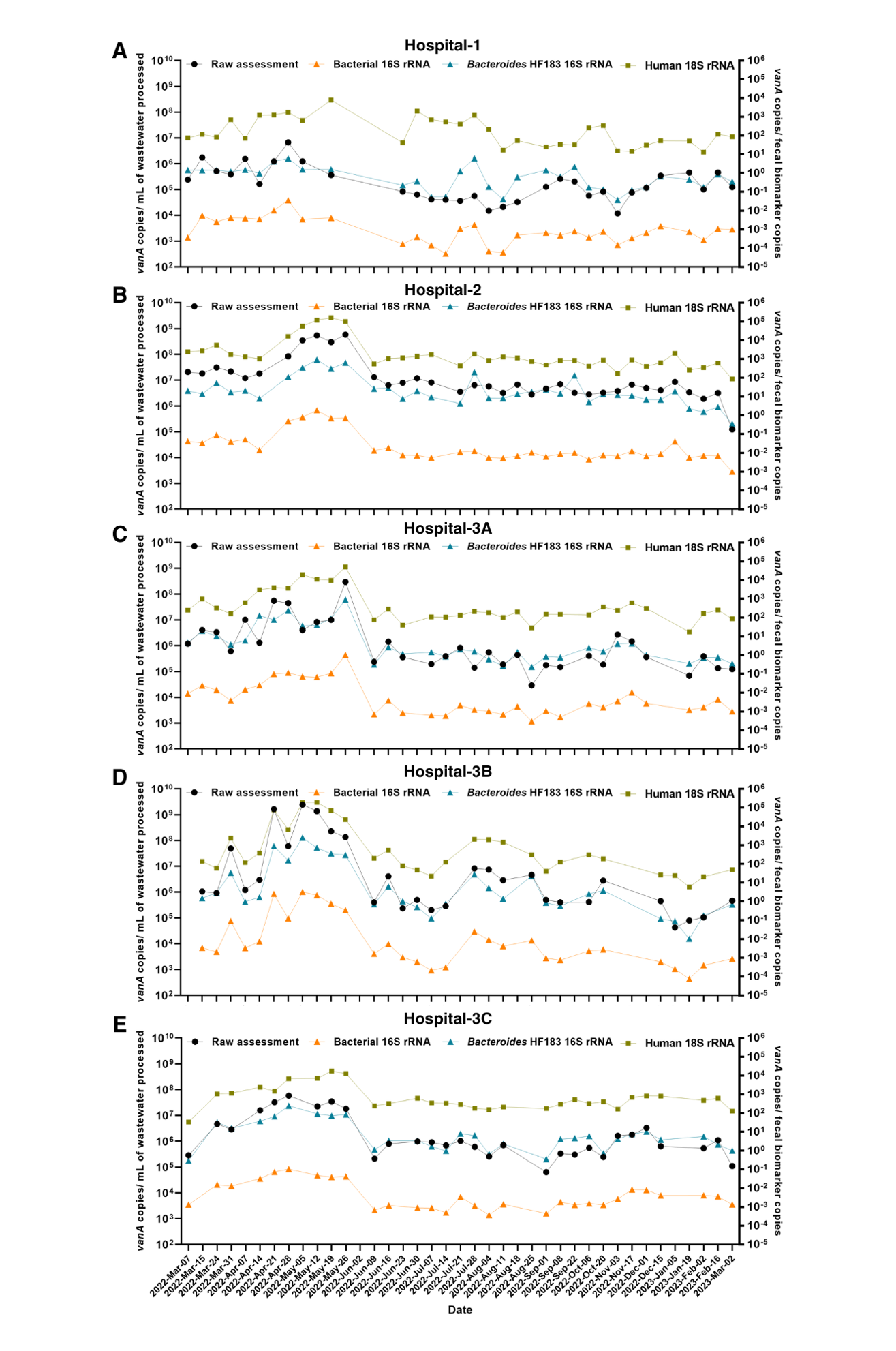

**Supplementary Figure 7. Normalization of *vanA* with different fecal biomarkers yields similar trends with longitudinal assessment.** Mean *vanA* quantities assessed as raw values (copies per mL of wastewater) and ratios of different fecal biomarkers (total bacterial 16S rRNA, *Bacteroides* HF183 16S rRNA, and human 18S rRNA) as measured in Calgary tertiary care hospitals by qPCR over time from March 7, 2022 to March 2, 2023.

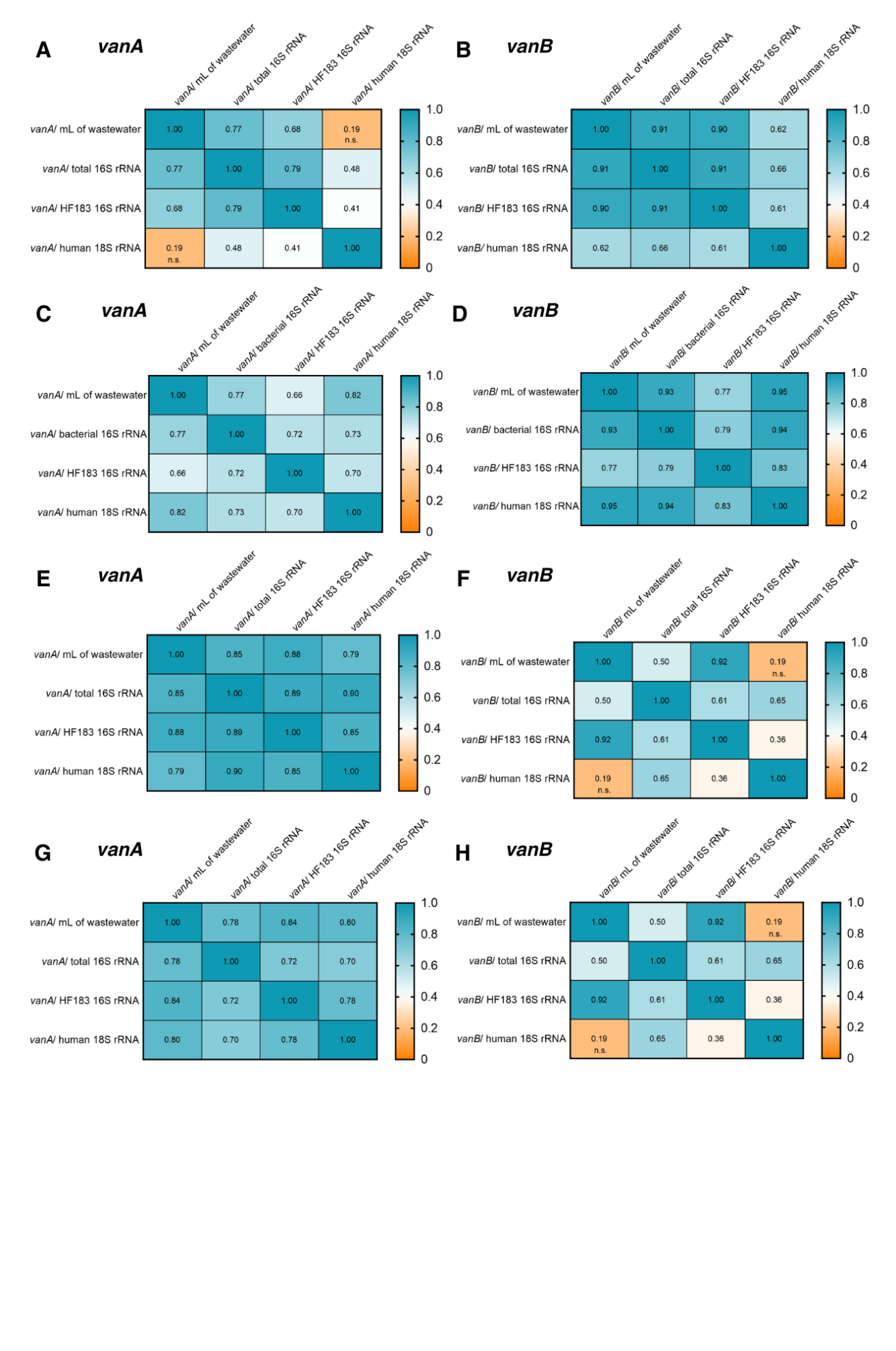

**Supplementary Figure 8. Comparison of raw assessment and fecal biomarker-normalized values correlated well with each other in the remaining adult hospital sites.** Spearman r correlations of *vanA* and *vanB* abundances detected in Hospital-1 (A-B), Hospital 2 (C-D), Hospital-3A (E-F) and Hospital-3B (G-H) wastewater assessed as raw (copies per mL of wastewater) and normalized with three fecal biomarkers – total bacterial 16S rRNA, *Bacteroides* HF183 16S rRNA, and human 18S rRNA). All Spearman r values were significant (p<0.05) unless otherwise noted – n.s. = not significant.

**Supplementary Table 4. Clinical isolates isolated from patients across the City of Calgary and Calgary-area hospitals.**

| **Site** | ***Enterococcus faecalis*** | | | ***Enterococcus faecium*** | | | **Comparator organisms** | |
| --- | --- | --- | --- | --- | --- | --- | --- | --- |
|  | **Total** | **Sensitive** | **VRE** | **Total** | **Sensitive** | **VRE** | ***E. coli*** | ***K. pneumoniae*** |
| **Calgary** | 3422 | 3422 (100%) | 0 | 126 | 113 (90%) | 13 (10%) | 17 855 | 2172 |
| **Hospital-1** | 191 | 189 (99%) | 2 (1%) | 57 | 41 (72%) | 16 (28%) | 304 | 90 |
| **Hospital-2** | 129 | 129 (100%) | 0 | 54 | 40 (74%) | 14 (26%) | 241 | 68 |
| **Hospital-3** | 421 | 421 (100%) | 0 | 135 | 100 (74%) | 35 (26%) | 749 | 232 |
| **Hospital-4 (pediatric)** | 37 | 37 (100%) | 0 | NA | NA | NA | 45 | NA |

**Supplementary Table 5. Recovery of VRE in clinical specimens in relation to reference organisms across multiple sites**

| **Site** | ***Ratio of Enteric pathogen to VRE*** | | | |
| --- | --- | --- | --- | --- |
|  | ***Escherichia coli*** | | ***Klebsiella pneumoniae* complex** | |
|  | **VRE : *E. coli* ratio** | **Rate at site relative to City overall** | **VRE : *K. pneumoniae* ratio** | **Rate at site relative to City overall** |
| **Calgary** | 0.0007 | Reference | 0.0060 | Reference |
| **Hospital-1** | 0.0526 | 72.2874 | 0.1778 | 29.7026 |
| **Hospital-2** | 0.0581 | 79.7861 | 0.2059 | 34.3982 |
| **Hospital-3** | 0.0467 | 64.1804 | 0.1509 | 25.2056 |
| **Hospital-4 (pediatric)** | NA | NA | NA | NA |

NA=data not available because of too few isolates collected to be reported.

**Supplementary Table 6.** Monthly average total vancomycin defined daily dose per hospital.

| **Month** | **Average Vancomycin Defined Daily Dose** | | |
| --- | --- | --- | --- |
|  | **Hospital-1** | **Hospital-2** | **Hospital-3** |
| March 2022 | 11.636 | 23.528 | 42.941 |
| April 2022 | 12.663 | 21.400 | 36.615 |
| May 2022 | 14.805 | 24.365 | 40.459 |
| June 2022 | 13.263 | 25.600 | 43.463 |
| July 2022 | 13.590 | 21.357 | 37.222 |
| August 2022 | 12.332 | 21.119 | 42.207 |
| September 2022 | 13.834 | 19.615 | 39.157 |
| October 2022 | 15.328 | 24.796 | 41.456 |
| November 2022 | 17.797 | 27.416 | 49.799 |
| December 2022 | 14.836 | 28.260 | 52.581 |
| January 2023 | 17.038 | 31.162 | 44.824 |
| February 2023 | 16.677 | 28.150 | 47.769 |

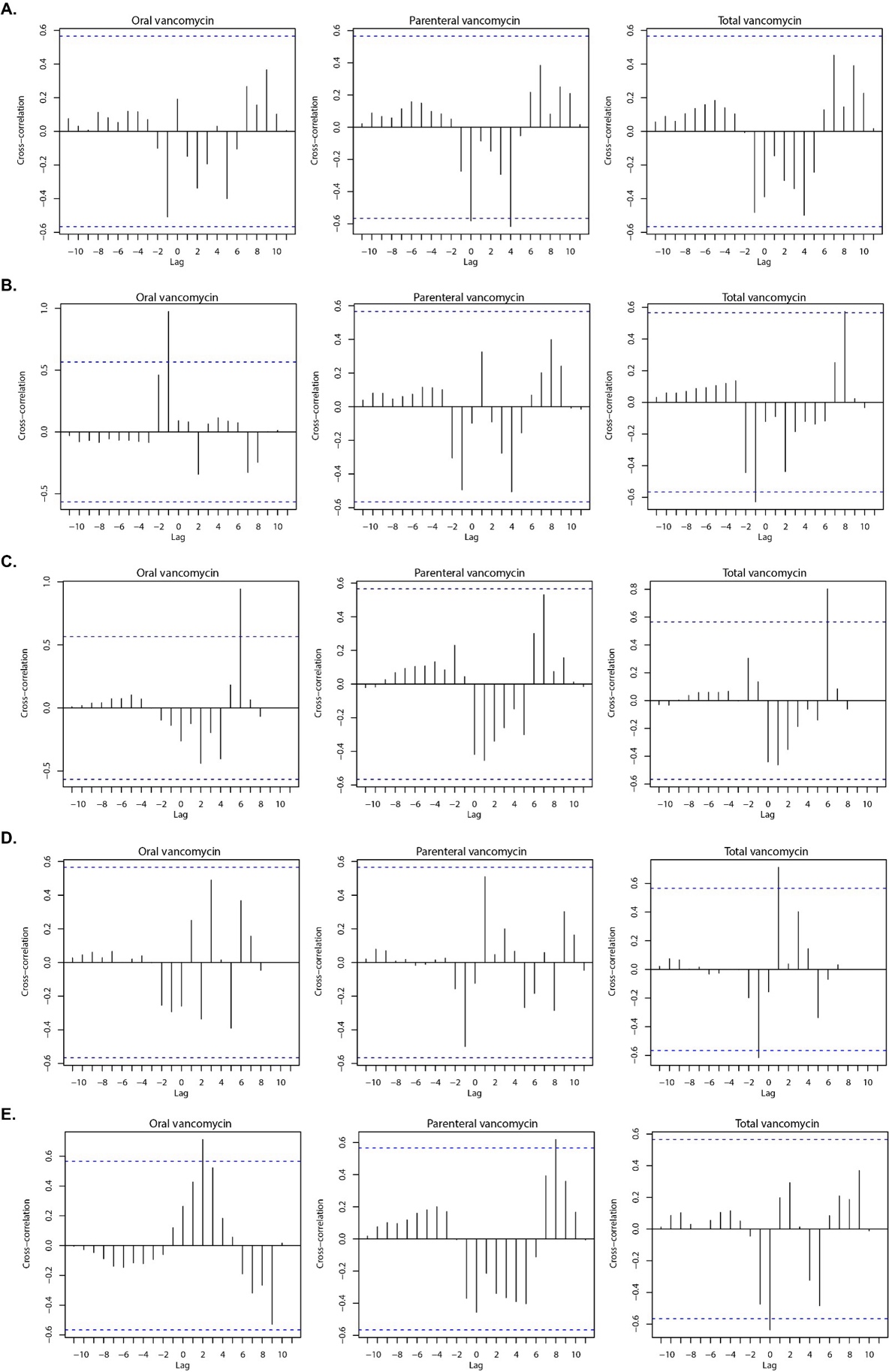

**Supplementary Figure 9. Comparative analysis of *vanA* concentrations and the monthly vancomycin prescribing data in adult hospitals.** Cross-correlation analysis of monthly oral, parenteral and total vancomycin prescribing (DDD/100 PD) with monthly averages of 16S rRNA-normalized *vanA* abundance in (A) Hospital-1, (B) Hospital-2, (C) Hospital-3A, (D) Hospital-3B, and (E) Hospital-3C from March 2022 to February 2023. Blue dotted lines denote the limits that determine the significance of the cross-correlation coefficients (95% confidence level).

**References**

1.	He Y-H, Ruan G-J, Hao H, Xue F, Ma Y-K, Zhu S-N, et al. Real-time PCR for the rapid detection of vanA, vanB and vanM genes. Journal of Microbiology, Immunology and Infection. 2020;53(5):746-50.

2.	Ritalahti KM, Amos BK, Sung Y, Wu Q, Koenigsberg SS, Löffler FE. Quantitative PCR targeting 16S rRNA and reductive dehalogenase genes simultaneously monitors multiple Dehalococcoides strains. Appl Environ Microbiol. 2006;72(4):2765-74.

3.	Acosta N, Bautista MA, Waddell BJ, McCalder J, Beaudet AB, Man L, et al. Longitudinal SARS-CoV-2 RNA wastewater monitoring across a range of scales correlates with total and regional COVID-19 burden in a well-defined urban population. Water Research. 2022;220:118611.

4.	Acosta N, Bautista MA, Waddell BJ, Du K, McCalder J, Pradhan P, et al. Surveillance for SARS-CoV-2 and its variants in wastewater of tertiary care hospitals correlates with increasing case burden and outbreaks. Journal of Medical Virology. 2023;95(2).

5.	AlbertaHealth. Respiratory virus dashboard Alberta, Canada: Alberta Health; 2023 [Available from: <https://www.alberta.ca/stats/dashboard/respiratory-virus-dashboard.htm>.

6.	Nadkarni MA, Martin FE, Jacques NA, Hunter N. Determination of bacterial load by real-time PCR using a broad-range (universal) probe and primers set. Microbiology (Reading). 2002;148(Pt 1):257-66.

7.	Greenwald HD, Kennedy LC, Hinkle A, Whitney ON, Fan VB, Crits-Christoph A, et al. Tools for interpretation of wastewater SARS-CoV-2 temporal and spatial trends demonstrated with data collected in the San Francisco Bay Area. Water Research X. 2021;12:100111.
